# Do sex hormones confound or mediate the effect of chronotype on breast and prostate cancer? A Mendelian randomization study

**DOI:** 10.1101/2021.04.20.21255783

**Authors:** Bryony Hayes, Tim Robinson, Siddhartha Kar, Katherine S Ruth, Konstantinos K Tsilidis, Tim Frayling, Anna Murray, Richard M Martin, Deborah A Lawlor, Rebecca C Richmond

**Author notes:** AUTHOR CONTRIBUTIONS BH and RCR conceived the study and BH carried out the data curation and analysis. The initial manuscript was drafted by BH, TR, RMM, DAL and RCR. All authors contributed to the interpretation of the results and critical revision of the manuscript.

## Abstract

**Background:** Previous research has demonstrated that a morning-preference chronotype is protective against both breast and prostate cancer. Sex hormones have been implicated in relation to both chronotype and the development of both cancers. This study aims to assess whether sex hormones confound or mediate the effect of chronotype on breast and prostate cancer risk using a Mendelian Randomization (MR) framework.

**Methods:** We obtained genetic variants strongly (p<5×10^−8^) associated with chronotype and sex hormones (total testosterone, bioavailable testosterone, sex hormone binding globulin (SHBG), and oestradiol from previously published genome-wide association studies (GWAS) that had been undertaken in UK Biobank and 23andMe (n≤244,207 females and n≤205,527 males). These variants were used to investigate causal relationships with risk of breast and prostate cancer using summary data from the largest available consortia in breast (nCases/nControls=133,384/ 113,789) and prostate cancer (nCases/nControls=79,148/61,106). This was achieved using a series of MR approaches: univariable, bidirectional and multivariable.

**Results:** Overall, we found evidence for a protective effect of genetically predicted tendency towards morning preference on both breast (OR=0.93, 95% CI:0.88, 1.00) and prostate (OR=0.90, 95% CI:0.83, 0.97) cancer risk. There was evidence that an increased tendency to morning preference reduces bioavailable testosterone levels in both females (mean SD difference=-0.08, 95% CI:-0.12, - 0.05) and males (mean SD difference=-0.06, 95% CI:-0.09, -0.03), and reduces total testosterone levels in females (mean SD difference=-0.07, 95% CI:-0.10, -0.03). We also found evidence to support higher total and bioavailable testosterone increasing the risk of breast cancer (OR=1.15, 95% CI:1.07, 1.23, OR=1.10, 95% CI:1.01, 1.19 respectively) and higher bioavailable testosterone increasing prostate cancer risk (OR=1.22, 95% CI:1.08, 1.37). While findings from univariable and bidirectional MR analyses indicated that testosterone may lie on the causal pathway between chronotype and cancer risk, there was evidence for a bidirectional association between chronotype and testosterone in females, implicating testosterone as both a confounder and mediator of the chronotype effect on breast cancer risk. However, the effects of chronotype remained largely unchanged when accounting for testosterone in multivariable MR, suggesting that any confounding or mediating effect is likely to be minimal.

**Conclusions:** This study has extended previous findings regarding the protective effect of chronotype on breast cancer and found evidence to suggest that morning preference also reduces prostate cancer risk in men. While testosterone levels were found to be closely linked with both chronotype and cancer risk, there was inconsistent evidence for the role of testosterone in mediating the effect of morning preference chronotype on both breast and prostate cancer. Findings regarding the potential protective effect of chronotype on both breast and prostate cancer risk are clinically interesting. However, this may not serve as a direct target for intervention, since it is difficult to modify someone’s morning/evening preference. Given this, further studies are needed to investigate the mechanisms underlying this effect and to identify other potential modifiable intermediates.

## INTRODUCTION

Breast and prostate cancers are hormonally related tumours with a significant burden of morbidity and mortality: an estimated 2.1 million women and 1.3 million men were diagnosed with these conditions in 2018(1). Despite improvements in cancer screening, treatment(2,3)and monitoring (4,5), they remain the most common cancers in women and men, respectively, and their incidence is increasing(6). Improved understanding of the aetiology of these cancers is an important step towards the development of effective primary prevention strategies.

Circadian rhythms underlying the sleep-wake cycle are implicated in cancer development, particularly in breast and prostate cancers(7,8). Variation in sleep-wake cycles between individuals(9) can manifest as circadian preference (chronotype), with most people fitting into one of three broad categories: morning-preference; evening-preference; or no preference. Using multivariable regression, we showed that a per category increase from extreme evening preference to extreme morning-preference chronotype was associated with a reduction (OR=0.95 (95% CI: 0.93, 0.98)) in breast cancer incidence in the UK Biobank prospective cohort study (n= 151,421; 2,732 incident cases(10)), and this result was consistent when triangulated across different methods, including Mendelian randomization (MR) (see below). A prostate case-control study (1095 cases and 1388 controls) found limited evidence for a difference in chronotype between cases and controls. However, among men with an evening preference, overall risk was higher among night shift workers compared with non-shift workers (OR 1.50; 95% CI 0.85, 2.66), whereas among men with no preference chronotypes, there was no difference in risk between night shift workers and non-shift workers (OR 1.02; 0.72, 1.44).(11).

An individual’s chronotype is largely determined by genetic variants(12), external environmental cues (e.g. light exposure(13)), and age(14). Typically, children exhibit morning-preference, which becomes progressively later during adolescence, then as adults age there is a gradual shift back towards morning preference(15). Generally, females reach these markers earlier than males, and males are more likely to retain their adolescent evening-preference into adulthood(9). However, sex differences reduce around 50 years of age, coinciding with the onset of menopause(15). This variability over time and by sex suggests that an individual’s chronotype may, in part, be hormonally driven.

Hormones, such as cortisol, melatonin and some sex steroids, have previously been studied in relation to chronotype(16–20). In particular, evening chronotype has been associated with higher levels of testosterone in males, even after adjusting for age(19),, with little evidence of an association of progesterone, dehydroepiandrosterone (DHEA) or testosterone with chronotype in females(20). However, in cross-sectional studies such as these, it is difficult to fully account for unobserved confounding and the direction of effect between chronotype and hormone levels is difficult to resolve.

MR is an approach which uses genetic variants robustly associated with exposures to estimate potential causal effects on outcomes. It attempts to overcome some potential limitations of conventional analyses used in observational studies, including confounding and reverse causation, though has additional potential biases(21–23). Using genetic variants related to chronotype and sex hormones, this approach may be used to investigate the bi-directional relationship between these traits and also to provide additional evidence regarding their effects on breast and prostate cancer. The statistical power and precision of MR analyses can be increased by using two-sample MR, in which summary genetic association data from independent samples representing genetic variant-exposure and genetic variant-outcome associations are combined in order to estimate causal effects(24).

Using two-sample MR, recent studies have reported a 7% reduction in breast cancer risk(25) and a 9% reduction in prostate cancer risk(26) per 25 and 30nmol/L increase in SHBG, respectively. A 15% increase in breast cancer risk and 22% increase in prostate cancer risk per 4nmol/L increase in bioavailable testosterone has also been reported(26). While MR studies for the effects of oestrogen on cancer risk in males and premenopausal females is lacking, a recent randomised control trial found that postmenopausal females undergoing oestrogen-only hormone replacement therapy (HRT) for 1 - 14 years experienced a 17 – 33% increase in breast cancer risk compared to never-users(27). A retrospective cohort study in post-menopausal females compared oestradiol levels from highest fifth to lowest (measured by immunoassay) and found an increased risk of breast cancer (OR=2.66; 95% CI: 1.99-3.54), with similar results reported across different oestradiol-measurement methods(28).

Regarding the effect of chronotype on breast cancer, in our previous study, we used results from both one-sample and two-sample MR analysis to support the multivariable regression result of a protective causal effect of morning chronotype on breast cancer risk (OR=0.85; 95% CI: 0.70, 1.03 and OR=0.88; 95% CI: 0.82, 0.93 per increased category towards morning preference, for one- and two-sample MR, respectively)(10).

The aims of this study were to use two-sample MR to explore whether: (i) the causal effect of chronotype on breast cancer risk found previously(10) remains consistent using newly available breast cancer data, including data on subtypes (29); (ii) chronotype exerts a causal effect on prostate cancer risk; and (iii) the effect of chronotype on breast, and any effect on prostate cancer risk, is mediated or confounded by sex hormones (total testosterone, bioavailable testosterone, sex hormone binding globulin (SHBG) and oestradiol).

## METHODS

### Overall Study Design Strategy

Our overall strategy was to perform a series of two-sample MR analyses. First, univariable MR (uvMR) was used to estimate the effects of: chronotype on breast and prostate cancer risk (step 1, figure 1A); and sex hormone levels on cancer risk (step 2, figure 1B). In step 3, bidirectional MR (bdMR) was used to determine the direction of associations between sleep traits and sex hormones (figure 1C)(22). In step 4, multivariable MR (mvMR) was used to further explore whether the chronotype effect on breast or prostate cancer is mediated by any of the hormones (figure 1D)(30,31). Only those sex hormones with a potential role in mediating the effect of chronotype on cancer risk in step 3 were used in subsequent mvMR. A potential mediating role was determined based on the following criteria: (i) evidence of a causal effect of chronotype on a specific cancer; (ii) evidence of a causal effect of chronotype on the specific sex hormone; (iii) evidence of a causal effect of the specific sex hormone on the specific cancer, and (iv) no evidence of a strong bidirectional effect. For further information regarding study design see figure 2 and supplementary methods 1.

**Figure 1.**
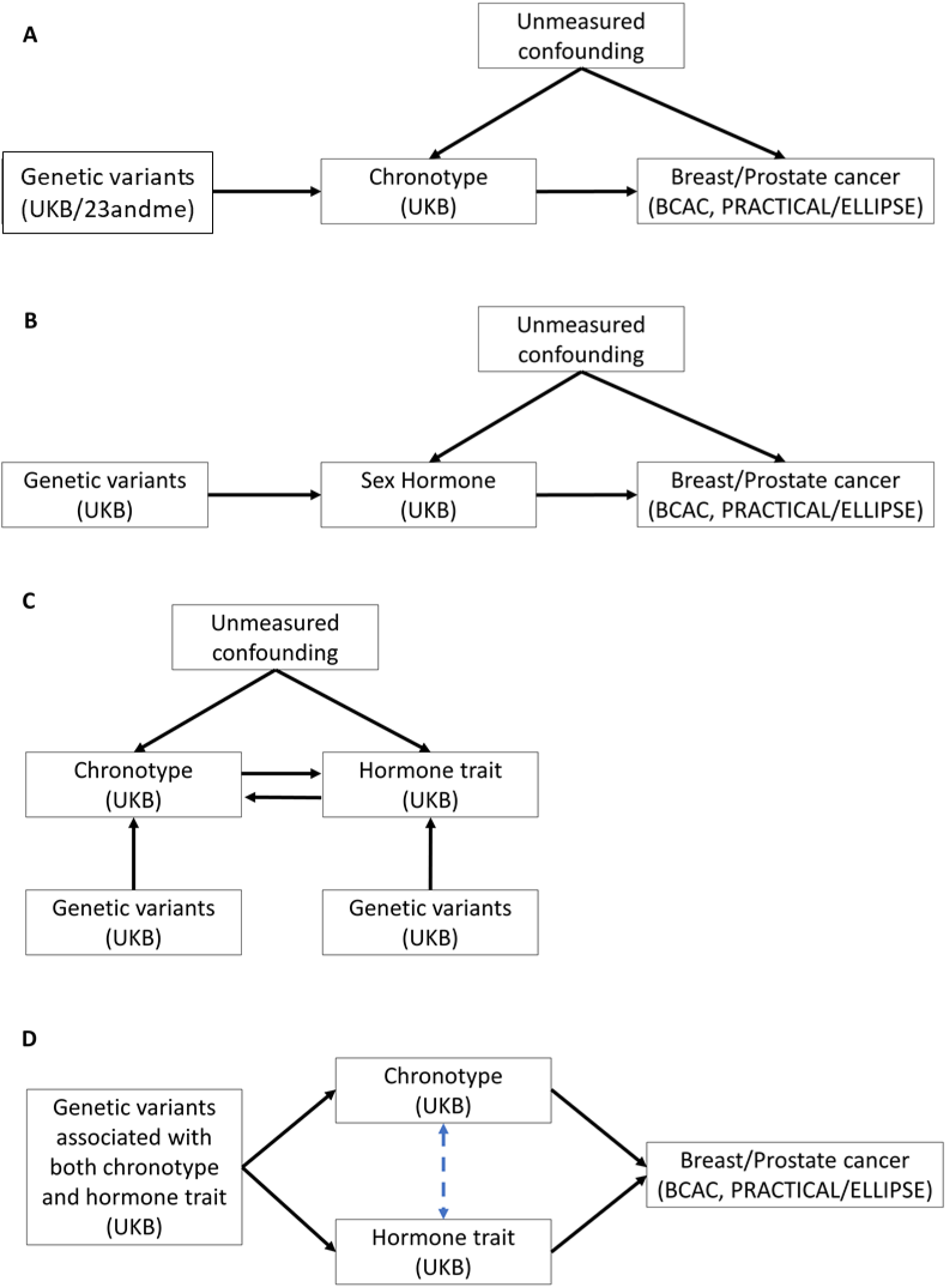
Schematics of MR analyses performed in this study. Participants datasets used to conduct GWAS’ given in parenthesis. A) Two-sample MR to determine the effect of chronotype on breast/prostate cancer. B) Two-sample MR to determine the effect of sex hormones (total testosterone, bioavailable testosterone, SHBG and oestradiol) on breast/prostate cancer. C) Bidirectional MR, used to identify directionality of relationship between chronotype and sex hormone traits. D) Multivariable MR to determine the direct effect of each exposure on breast/prostate cancer risk. The blue dashed arrow in this schematic depicts adjustment of one exposure effect in the presence of the other.

**Figure 2:**
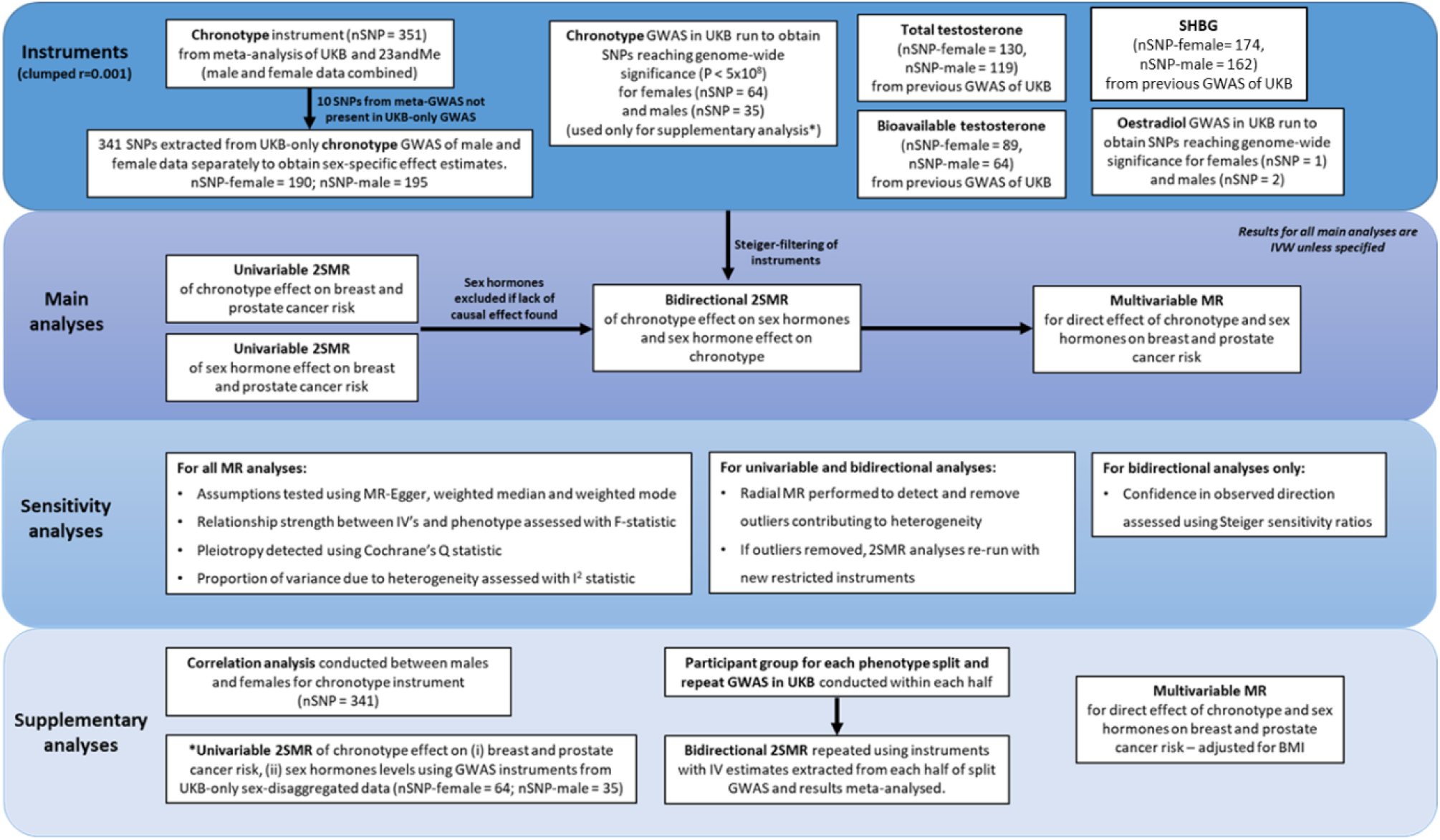
Overview of instruments and all main and supplementary analyses.

### Genome-wide association study populations

#### UK Biobank

Summary genetic data for chronotype and sex hormones were obtained from GWAS conducted in the UK Biobank (UKB). More details on the study, the genotyping procedure, and the data sources for chronotype and the sex hormones are described in supplementary methods.

While GWAS summary statistics for both chronotype and sex hormones using data from UK Biobank currently exist(26,32), we conducted our own GWAS for chronotype and sex hormones separately in men and women, given the sex-specific nature of the cancers we were investigating. For this, we used a linear mixed model (LMM) association method to account for relatedness and population stratification, as implemented in BOLT-LMM (v2.3)(33). More details of this approach can be found in supplementary methods 2.

#### Breast Cancer Association Consortium and Prostate Cancer Association Group

For breast and prostate cancer outcomes, genome-wide association studies (GWAS) summary data were obtained from the Breast Cancer Association Consortium (BCAC)(34),(29); and the Elucidating Loci Involved in Prostate Cancer (ELLIPSE) and Prostate Cancer Association Group to Investigate Cancer Associated Alterations in the Genome (PRACTICAL) consortia(35,36) (PRACTICAL/ELLIPSE). Sample sizes for these GWAS are shown in Table 1.

**Table 1.** Sample sizes for exposure and outcome GWAS’ used in this study for male and female analyses. Number of SNPs (nSNP) given for clumped instruments from uvMR analyses.

| Exposures | Female | Male | nSNP (F/M) | Study |
| --- | --- | --- | --- | --- |
| Chronotype | 244,207 | 205,527 | 190 / 195 | UKB |
| Total Testosterone | 199,596 | 200,159 | 130 / 119 |  |
| Bioavailable Testosterone | 180,386 | 184,205 | 89 / 64 |  |
| SHBG | 214,989 | 185,221 | 174 / 162 |  |
| Oestradiol | 53,391 | 17,134 | 1 / 2 |  |
| Outcomes | Cases | Controls |  |  |
| Breast cancer (overall) | 133,384 | 113,789 | - | BCAC |
| Breast cancer (subtypes) | 106,491 | 94,407 | - |  |
| Prostate cancer (overall) | 79,148 | 61,106 | - | ELLIPSE/PRACTICAL |

The GWAS of overall breast cancer included 133,384 cases and 113,789 controls from 67 studies, using data from OncoArray and iCOGS arrays which were meta-analysed(34). Summary statistics were also obtained on 106,491 cases on whom breast cancer subtype data was available: Luminal A (oestrogen receptor positive (ER+)/progesterone receptor positive (PR+), human epidermal growth factor receptor 2 (HER2)); Luminal B (ER+/PR+/-, HER2+); Luminal B HER2 negative (ER+/PR+/-, HER2-, Ki-67 > 14%); HER2 (ER-, PR-, HER2+); and Triple Negative (ER-, PR-, HER2-), as well as 94,407 controls(29). All participants were females of European ancestry(34),(29). Details of genotyping and imputation for BCAC can be found in supplementary methods 3.

The GWAS for prostate cancer included data from the PRACTICAL (46,939 cases and 27,910 controls from 52 studies) and ELLIPSE (34,379 cases, 33,164 controls from 78 studies) consortia(35,36). The resulting overall meta-analysis included 79,148 cases and 61,106 disease free controls. All participants included in these analyses were males of European ancestry. Details of genotyping and imputation for ELLIPSE/PRACTICAL can be found in supplementary methods 4.

### Constructing Genetic instruments

We next identified genetic instruments for chronotype and sex hormones, for which SNP-exposure estimates were obtained from the sex-specific GWAS for chronotype and sex hormones in UK Biobank.

For chronotype, we identified two genetic instruments. The first comprised SNPs identified in the largest published GWAS of meta-analysed women and men combined from UKB and 23andme (N = 697,828) (37). Of the 351 single nucleotide polymorphisms (SNPs) that were genome-wide significant in that study, we identified 341 of those SNPs in UKB and extracted association summary results (SNP, effect allele, other allele, beta (increase in chronotype category per effect allele), SE and p-value) separately in females and males. Those statistics were then used in our two-sample MR (irrespective of whether they reached genome-wide significance within the sex-specific GWAS). This approach maximises statistical power but might lack specificity for each sex. In the second approach we only used summary results (as above) for SNPs that reached GWAS significance (p < 5×10^−8^) in the sex specific GWAS within UKB. This will have less statistical power and may be more prone to weak instrument bias but may be a more relevant sex-specific instrument. Following LD-clumping, to obtain independent SNPs for our genetic instrument (using a threshold of R^2^ = 0.001), there were 190 and 195 SNPs for chronotype in females and males, respectively for the first approach and 54 and 32 SNPs, respectively in the second approach. Supplementary tables 1-2 provides details of the 341 SNPs identified in the published GWAS that were present in our UKB GWAS, the 190 and 195 used as genetic instruments in our first approach and the 54 and 32 used in our second approach.

Sex specific genetic instruments for total testosterone, bioavailable testosterone and SHBG were obtained from the previously published GWAS’ conducted in UKB(26). The GWAS for total testosterone for women (N = 230,454) and men (N = 194,453) identified 254 and 231 SNPs respectively that were statistically independent, which LD-clumping then restricted to 130 and 119 SNPs, respectively. The GWAS for bioavailable testosterone for women (N = 188,507) and men (N = 178,782) identified 180 and 125 SNPs respectively that were statistically independent, which LD-clumping then restricted to 89 and 64 SNPs, respectively. The GWAS for SHBG for women (N = 189,473) and men (N = 180,726) identified 359 and 357 SNPs respectively that were statistically independent, which LD-clumping then restricted to 192 and 177 SNPs, respectively. Instruments for oestradiol were obtained from GWAS we undertook in UKB separately for females (N = 53,391) and males (N = 17,134), which identified 40 and 378 SNPs respectively that were genome wide significant (GWS), resulting in 1 and 2 SNPs following LD-clumping for females and males, respectively. A high proportion of the oestradiol assay results had values below the limit of detection (80%) and were excluded from the GWAS(38).

The number of SNPs used to instrument chronotype and each of the sex hormone measures can be found in table 1. The summary statistics for all instruments used in this study are available in supplementary data (supplementary tables 1-2).

### Statistical Analyses

#### Two-Sample Mendelian Randomization

For uvMR, bdMR and mvMR analysis, the TwoSampleMR package was used to combine and harmonize genetic summary data for chronotype, sex hormones and breast and prostate cancer. The inverse variance weighted (IVW) approach was used for the main analysis, whereby a causal effect is estimated from the slope of a regression line through the weighted SNP-mean exposure vs SNP-mean outcome associations (orientated to be positive) with the line constrained to have an intercept of zero. Since only 1 SNP contributed to the female oestradiol instrument, MR results were instead calculated using a Wald ratio(39).

#### Assessment of MR Assumptions

MR analysis relies on three key assumptions: i) IVs must be robustly associated with the exposure of interest; ii) IVs must be independent of confounders of the exposure-outcome association; iii) IVs must only influence the outcome through the exposure of interest(40).

We conducted sensitivity analyses to test MR assumptions. Univariable and conditional F-statistics were calculated to assess the strength of the relationship between the IVs and phenotype(41),(42)(43). To test the assumption of no unbalanced horizontal pleiotropy we conducted MR-Egger(44), weighted median(45) and weighted mode(46) MR (see supplementary methods 5). Scatter plots were used to visualise consistency between IVW, MR-Egger, median and mode effect estimates. To further test the assumption of no horizontal pleiotropy, we explored between SNP heterogeneity using Cochran’s Q and leave one out analyses, as presence of heterogeneity may be due to pleiotropy of one or more SNPs(22),(47). I^2^ statistics were used to estimate the proportion of the variance between IV estimates that is due to heterogeneity(48). Radial-MR was also conducted to identify IVs with the largest contribution to heterogeneity (alpha = 0.05) and mark them as outliers(49). To assess their impact, these outliers were removed from the instrument and the data is then reanalysed. Steiger filtering was applied to instruments in the bdMR analysis of chronotype and sex hormones to reduce the likelihood of erroneous results due to pleiotropy (here where the SNP has a primary influence on the outcome rather than the exposure) (50). Steiger sensitivity ratios (R) were calculated to ensure the likelihood that the observed direction of association was correct(51). Since only 1 and 2 SNPs contributed to female and male oestradiol instruments, it was not possible to conduct any sensitivity analyses for oestradiol.

To accommodate additional two-sample MR assumptions, we restricted analysis to European-only individuals in all datasets and ensured sex-specific GWAS were used to meet the assumption that the exposure and outcome samples come from the same underlying population. We also performed harmonization of the direct of effects between the SNPs in the exposure and outcome GWAS. Palindromic SNPs were harmonized if they were aligned and the minor allele was <0.3, otherwise they were excluded.

While sample overlap between UKB, BCAC and PRACTICAL/ELLIPSE is negligible, for the bdMR analysis, genetic estimates for both chronotype and sex hormones were determined in the same sample (UKB), violating the assumption of independent samples in two-sample MR. A supplementary analysis was carried out to investigate the magnitude of bias caused by overlapping samples(52). To evaluate this, we performed split-sample MR analyses, where we randomly split UKB participants into two halves and performed additional GWAS for chronotype and the sex hormones in each of the subgroups. We used SNP-chronotype effect estimates obtained from the first subgroup (sample 1) and SNP-hormone effect estimates obtained from the second independent subgroup (sample 2), and performed IVW analysis. This method was repeated for the SNP-chronotype effect estimates obtained from sample 2 and SNP-hormone estimates from sample 1. The two MR estimates were then meta-analysed under a fixed-effects model to obtain an effect of chronotype on sex hormones. This approach was conducted separately for males and females.

#### Supplementary analyses

Supplementary analyses were conducted to evaluate the effects of chronotype and sex hormones on breast cancer stratified by subtype: Luminal A (ER+/PR+, HER2-); Luminal B (ER+/PR+/-, HER2+); Luminal B (ER+/PR+/-, HER2-, Ki 67 > 14%); HER2 (ER-, PR-, HER2+); and Triple Negative (ER-, PR-, HER2-). An I^2^ statistic was calculated to assess variability in causal estimates between breast cancer subtypes(53).

In addition, evidence from previous research suggests that adiposity is intrinsically linked with certain sleep traits(54,55), sex hormones(56–58), and both breast and prostate cancer(59). As such, supplementary mvMR analyses were conducted in which we adjusted the effect of chronotype on cancers for body mass index (BMI), and additionally for both BMI and sex hormone(s).For this, instrumental SNPs for BMI were obtained from a UK Biobank GWAS of males and females (N = 461,460) (60) and sex-specific association summary results for these SNPs (nSNP = 458) were obtained from female (N = 171,977) and male (N = 152,893) GIANT consortia GWAS’ (61), which LD-clumping then restricted to 336 and 334 SNPs, respectively.

MR analyses used the R packages “Two SampleMR”(60) and “MVMR”(62), R version 4.0.2.

#### Data Availability

Summary data for the GWAS conducted as part of this study have been made available on the OpenGWAS database (gwas.mrcieu.ac.uk)

This research was conducted using the UK Biobank Resource under application number 16391.

Summary data for the breast cancer GWAS used in this study is available at: http://bcac.ccge.medschl.cam.ac.uk/bcacdata/oncoarray/oncoarray-and-combined-summary-result/gwas-summary-associations-breast-cancer-risk-2020/

Summary data for the prostate cancer GWAS used in this study is available at: http://practical.icr.ac.uk/blog/?page_id=8164

## RESULTS

### Univariable MR: Effect of Chronotype on Breast and Prostate Cancer Risk

The correlation (r) of the effect estimates for the 341 SNPs for chronotype in males and females was 0.60 (supplementary figure 1). The genetic variants (190 SNPs) contributing to the main chronotype instrument in females had a combined r^2^ of 1.8% and an F-statistic of 22.75, and in males (190 SNPs) had an r^2^ of 1.7% and an F-statistic of 17.18 (supplementary table 3). There was low to moderate heterogeneity between SNPs used within the instruments: the I^2^ for MR of overall breast cancer was 41% and for overall prostate cancer was 26% (supplementary table 3).

We observed a reduction in risk of breast cancer per category increase in chronotype from extreme evening preference to extreme morning preference (OR=0.93, 95% CI:0.88, 1.00) (figure 3A). There was also evidence of a protective effect of morning preference on prostate cancer (OR=0.90, 95% CI:0.83, 0.97) (figure 3B). Results from MR-Egger, weighted median and weighted mode methods were similar to IVW estimates (Figure 3 and supplementary table 4). In Radial MR, three SNPs were identified as potential influencing outliers in the main IVW analyses of chronotype on breast cancer. When removed, the results were unchanged (OR=0.94, 95% CI:0.89, 1.00) (supplementary table 5). There were no outliers identified in the analysis of chronotype on prostate cancer.

**Figure 3:**
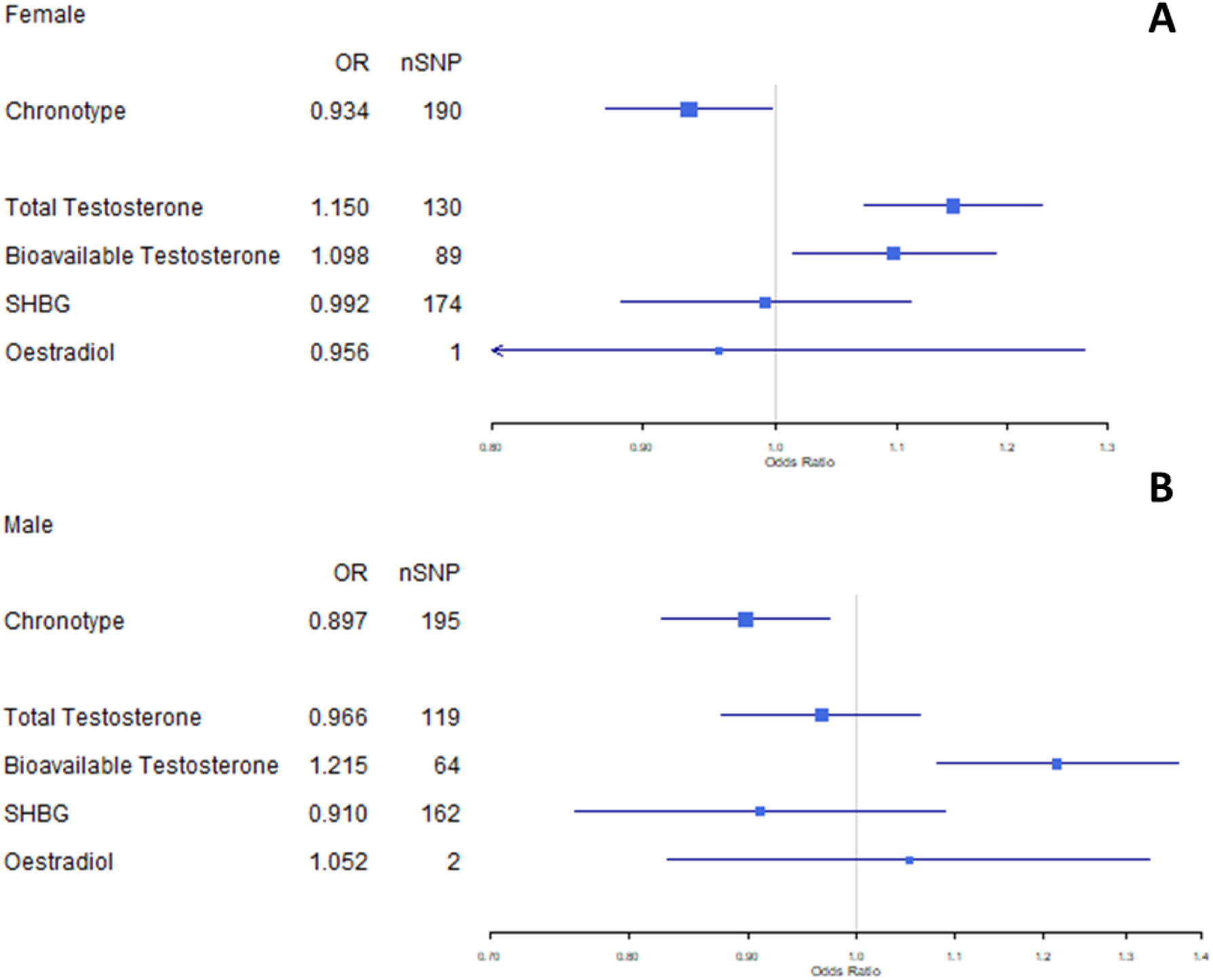
Forest plot of chronotype and sex hormone effect on brease (3A) and prostate (3B) cancer risk. All estimates calculated IVW, except for Oestradiol in femals, which was calculated using Wald ratio analysis.

The effect of chronotype on breast cancer was broadly similar between breast cancer subtypes, and heterogeneity for MR analyses between breast cancer subtypes was low (I^2^=26%) (supplementary figure 2).

Using UKB-only sex-specific instruments for chronotype, the results for chronotype on breast (OR=0.92, 95% CI: 0.83, 1.01) and prostate (OR=0.95, 95% CI: 0.81, 1.12) cancer were consistent with those in the main analysis, although less precisely estimated (supplementary table 6).

### Univariable MR: Effect of Sex Hormones on Breast and Prostate Cancer Risk

The genetic variants contributing to the instruments for total testosterone, bioavailable testosterone, SHBG and oestradiol in females had a combined r^2^ of 3.4%, 5.5%, 6.4% and 0.1%, respectively. F-statistics for these instruments are 44.48 – 83.59 (supplementary table 7). The genetic variants contributing to the instruments for total testosterone, bioavailable testosterone, SHBG and oestradiol in males had a combined r^2^ of 4.8%, 2.5%, 10.9% and 0.4%, respectively. F-statistics for these instruments are 36.75 – 143.35% (supplementary table 7).

Amongst females, an increased risk of breast cancer was observed with increasing total (OR=1.15, 95% CI: 1.07, 1.23 per SD) and bioavailable (OR=1.10, 95% CI: 1.01, 1.19 per SD) testosterone, but not in relation to SHBG (OR=0.99, 95% CI: 0.89, 1.11 per SD) (figure 3A). Amongst males, an increased risk of prostate cancer was observed for bioavailable testosterone (OR=1.22, 95% CI: 1.08, 1.37) but not in relation to total testosterone (OR=0.97, 95% CI: 0.86, 1.07), or SHBG (OR=0.91, 95% CI: 0.76, 1.09) (figure 3B). The effect of oestradiol on both breast (OR=0.96, 95% CI: 0.72, 1.27) and prostate (OR=1.05, 95% CI: 0.83, 1.33) cancers was imprecisely estimated due to insufficient SNPs, and therefore oestradiol has been excluded from subsequent bdMR and mvMR analyses. Given the lack of evidence for a causal effect on breast or prostate cancer risk, SHBG has been excluded from all further analyses, and total testosterone has been excluded from analyses of prostate cancer.

There was moderate to high heterogeneity between SNPs used within the sex hormone instruments (excluding oestradiol): the I^2^ for MR of sex hormones on overall breast cancer was 62 - 69% and for overall prostate cancer was 58 - 74% (supplementary table 7). However, results from MR-Egger, weighted median and weighted mode methods were similar to IVW estimates (Figure 3 and supplementary table 8). In Radial MR there were five, three and three SNPs identified as potential influential outliers in the main IVW analyses of total testosterone, bioavailable testosterone and SHBG on breast cancer, respectively. There were also eight, four and eight SNPs identified in the main IVW analyses of total testosterone, bioavailable testosterone and SHBG on prostate cancer, respectively. In all radial MR analyses, when outliers were removed the results were unchanged (supplementary table 9).

For MR of sex hormones stratified by breast cancer subtype (supplementary figure 3 and supplementary table 10), the increased risk reported for total and bioavailable testosterone on overall breast cancer was consistent with all luminal-type breast cancers. However, for total testosterone on triple negative breast cancer, estimates were consistent with a reduced risk (OR=0.91, 95% CI:0.81, 1.03) while there was little evidence for an effect on HER2-Enriched breast cancer (OR=0.98, 95% CI:0.81, 1.18). Findings were similar with respect to bioavailable testosterone and breast cancer subtypes. Although little evidence was found for an effect of SHBG on overall breast cancer, subtype-specific analyses found reduced risk of luminal-A (OR=0.90, 95% CI:0.78, 1.04) and luminal-B HER2-negative (OR=0.90, 95% CI:0.74, 1.11) breast cancers, and increased risk of luminal-B (OR=1.09, 95% CI:0.88, 1.35), HER2-Enriched (OR=1.28, 95% CI:0.95, 1.71) and triple negative (OR=1.19, 95% CI:0.99, 1.44) breast cancers, but these were imprecisely estimated.

### Bidirectional MR: Reciprocal Effects between Chronotype and Sex Hormones

The exposures used in the bdMR analyses underwent Steiger filtering. The removal of SNPs more strongly associated with the outcome were removed, resulting in greater confidence in the direction of association as determined by corresponding sensitivity ratios (supplementary table 11). In females, the variance (r^2^) explained by the Steiger-filtered chronotype instrument was 1.9% compared to 3.9%, and 3.8% for total testosterone and bioavailable testosterone respectively (supplementary table 11). This is comparable to the variance explained in males, where r^2^ for chronotype was 1.7%, compared with 3.0% for bioavailable testosterone (supplementary table 11).

Amongst females (figure 4A), bidirectional relationships were observed for chronotype and the testosterone measures. Morning preference was found to lower total testosterone (mean SD difference=-0.07, 95% CI:-0.10, -0.03 per category increase), and total testosterone in turn was inversely related to morning preference (difference in category=-0.06, 95% CI:-0.10, -0.03 per SD increase). Similarly, morning preference was found to lower bioavailable testosterone (mean SD difference=-0.08, 95% CI:-0.12, -0.05 per category increase), and bioavailable testosterone was inversely related to morning preference (difference in category=-0.04, 95% CI: -0.08, 0.00 per SD increase).

**Figure 4.**
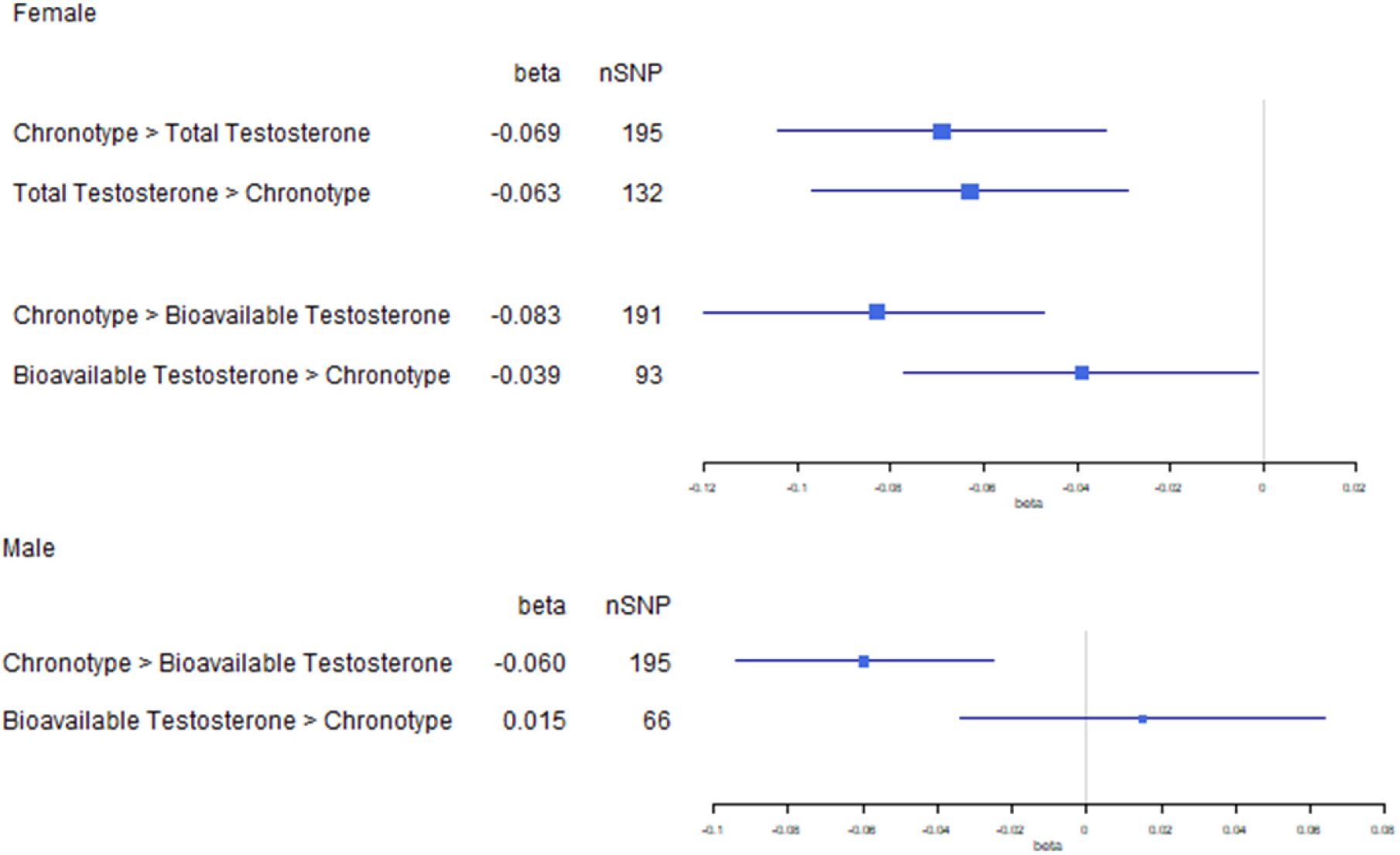
Forest plot of bdMR results for chronotype and sex hormones in female (4A) and male (4B) datasets. All instruments in these analyses have been subjected to Steiger-filtering.

Amongst males (figure 4B), morning preference was found to lower bioavailable testosterone (mean SD difference=-0.06, 95% CI:-0.09, -0.03 per category increase), but there was lack of corresponding evidence for an effect in the reverse direction (difference in mean category=0.02, 95% CI:-0.03, -0.06 per SD increase).

Overall, the bdMR analyses suggest that the relationship between chronotype and testosterone may be bidirectional for females, but with a prevailing effect from chronotype to testosterone in males (figure 4). Radial MR was performed for main analyses and bdMR was then repeated. The results of these analyses were consistent with the main results (supplementary tables 12). Bidirectional MR was also repeated using split-sample MR analysis and the results were largely consistent with those reported in the main analyses (supplementary figure 4).

### Multivariable MR: Direct Effects of Chronotype and Sex Hormones on Cancer Risk

Conditional F-statistics were calculated for all main mvMR analyses and varied from 7.08 to 18.36 (supplementary table 13).

In females (figure 5A), the reduced risk of breast cancer per category increase in chronotype observed with uvMR (OR=0.93, 95% CI: 0.88, 1.00) was largely unchanged when accounting for total testosterone (OR=0.93, 0.85-1.02) or bioavailable testosterone (OR=0.95, 95% CI: 0.88, 1.03). When accounting for chronotype, the increased risk of breast cancer observed per SD increase in total testosterone was consistent between uvMR (OR=1.15, 95% CI: 1.07, 1.23) and mvMR (OR=1.15, 95% CI: 1.04, 1.27) results. The increased risk of breast cancer observed per SD increase in bioavailable testosterone was also broadly consistent between uvMR (OR=1.10, 95% CI: 1.01, 1.19) and mvMR (OR=1.15, 95% CI: 1.03-1.28).

**Figure 5.**
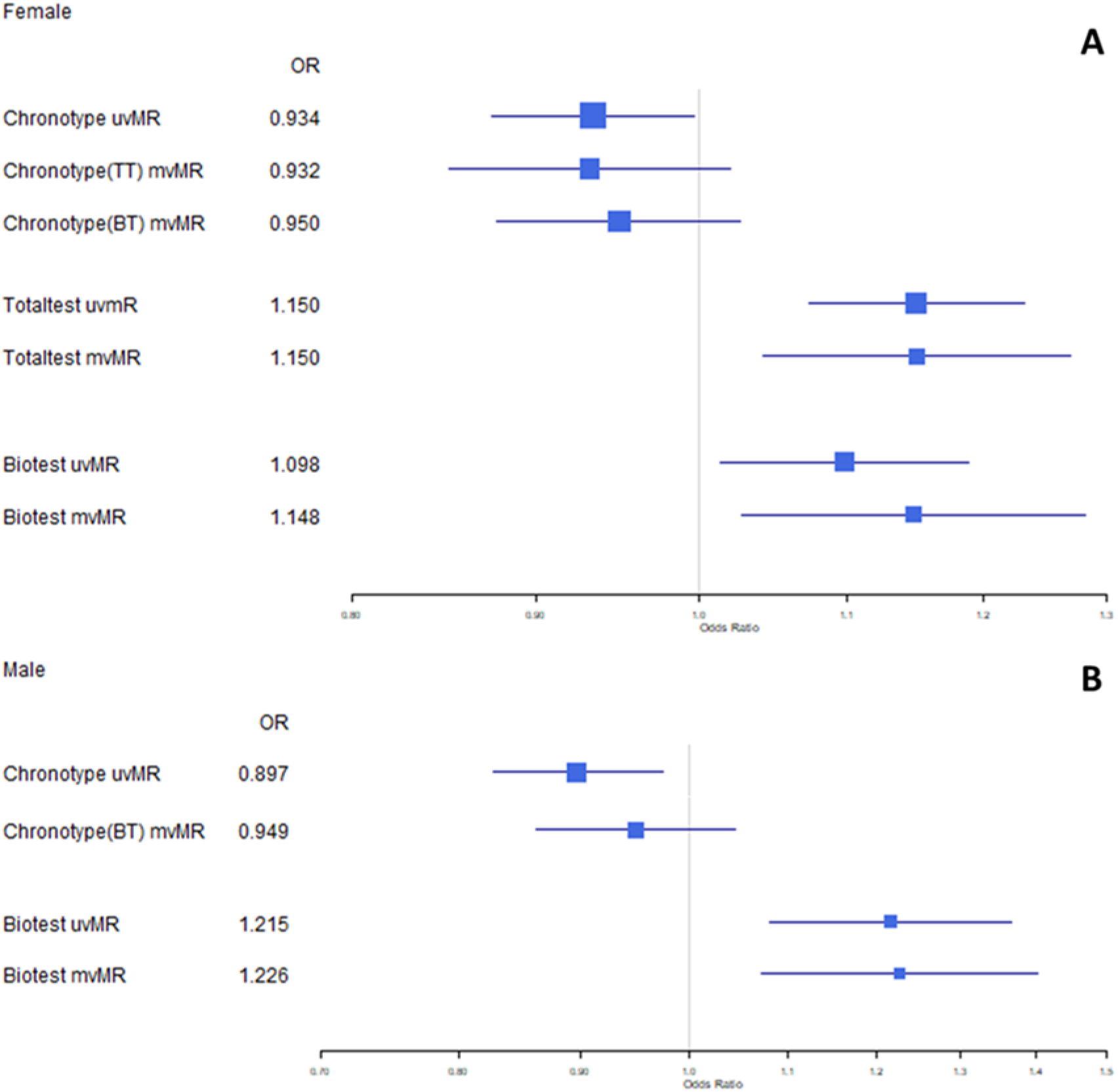
Forest plot of mvMR results for chronotype and sex hormones in female (5A) and male (5B) datasets. uvMR results provided for comparison.

In males (figure 5B), the reduced risk of prostate cancer per category increase in chronotype observed with uvMR (OR=0.90, 95% CI:0.83, 0.97) was broadly consistent when accounting for bioavailable testosterone (OR=0.95, 95% CI: 0.86, 1.05). Bioavailable testosterone also remained largely consistent between uvMR (OR=1.22, 95% CI: 1.08, 137) and mvMR results (OR=1.23, 95% CI: 1.08, 1.40) when accounting for chronotype.

Supplementary mvMR analyses were conducted to adjust for the effects of BMI (supplementary table 14). In females, (supplementary figure 5) the reduced risk of breast cancer per category increase in chronotype observed for uvMR was attenduated when adjusting for body mass index (BMI) (OR=1.00, 95% CI:0.85, 1.18). In mvMR, the direct effect of chronotype on breast cancer was directionally consistent when adjusting for bioavailable testosterone and BMI (OR=0.96, 95% CI:0.79, 1.17), and for total testosterone and BMI (OR= 0.91, 95% CI:0.73, 1.14). However, in all cases the results were imprecisely estimated. The direct effect of both total and bioavailable testosterone persisted with addition of BMI in the models. In males (supplementary figure 6), the direct effects obtained for chronotype and bioavailable testosterone were largely consistent with those found for uvMR.

## DISCUSSION

This study has assessed the evidence for the role of sex hormones in mediating or cofounding the effect of chronotype on risk of breast and prostate cancer using a series of MR analyses. Overall, we found evidence for a protective effect of genetically predicted tendency towards morning preference on both breast and prostate cancer risk. There was evidence that a tendency to morning preference reduces both total and bioavailable testosterone levels in males and females. We also found evidence to support higher testosterone increasing the risk of both breast and prostate cancer risk. The effect of testosterone on prostate cancer was more prominent when assessing bioavailable rather than total testosterone. While findings from univariable and bidirectional MR analyses indicated that testosterone may lie on the causal pathway between chronotype and cancer risk, there was evidence for a bidirectional association between chronotype and testosterone in females, suggesting that these sex hormones may have the capacity to both confound and mediate the chronotype effect on breast cancer risk. However, the effects of chronotype remained largely unchanged when accounting for testosterone in multivariable MR suggesting that any mediating effect is likely to be minimal. The effect of oestradiol on breast and prostate cancer was very imprecisely estimated and may have been influenced by weak instrument bias as we only had one SNP to instrument for oestradiol in females, and two SNPs in males. Having so few SNPs meant that we could not explore potential pleiotropy and therefore we did not explore this sex hormone further.

The effect of morning-preference on breast cancer risk is consistent, although more conservative, than our previously reported two-sample MR findings (OR=0.93, 95% CI:0.88, 1.00 vs OR 0.88, 95% CI:0.82, 0.93 per category increase in chronotype)(10). This may be attributed to differences in the genetic instruments used in the two analyses, with a more stringent threshold for obtaining independent SNPs in the current analysis (LD r^2^=0.001 vs 0.01) and the larger sample size of the breast cancer GWAS(29) (n= 247,173 vs n = 228,951). The chronotype effect on prostate cancer risk (OR=0.90, 95% CI:0.83, 0.97) was more pronounced in the current study than previously reported results obtained from logistic regression analyses, where there was a lack of association between morning and no preference (OR=0.98, 95% CI:0.79, 1.21) and a slightly higher risk for evening vs no preference (OR=1.10, 95% CI:0.82, 1.46), although confidence intervals are overlapping (11).

Our findings of increased testosterone increasing breast and prostate cancer risk are comparable to a previously published study (26) which also utilised MR to estimate effects of total (OR=1.14, 95% CI:1.08, 1.20) and bioavailable testosterone (OR=1.15, 95% CI: 1.05, 1.25) on breast cancer, and an effect of bioavailable testosterone on prostate cancer risk (OR=1.23, 95% CI: 1.13, 1.33), consistent with those reported in this study. Similarly, our findings that increasing SHBG reduces both breast and prostate cancer were consistent with previously reported MR results(25,26).

That morning-preference chronotype was associated with lower testosterone levels in males is in-keeping with previous cross-sectional studies(19,20). However, the finding that morning preference chronotype lowers testosterone levels in females is in contrast to previous null findings(20). This is likely due to the very small number of female participants (N=47) as many were excluded for testosterone levels below the limit of sensitivity. Furthermore, both prior studies inferred from association analyses that chronotype is dependent on testosterone levels, whereas bidirectional MR analyses in the currently study enabled us to orient the causal direction of this association, suggesting that testosterone levels are dependent on chronotype in males, and that there is a bidirectional relationship between chronotype with testosterone in females. A bidirectional relationship between these traits in females indicates that these hormones may serve as both confounders and mediators of the chronotype effect on breast cancer. However, results of the mvMR analyses did not support evidence for total testosterone or bioavailable testosterone acting as a mediator in males or females, since the effect of chronotype on breast and prostate cancer was found to be largely independent of these sex hormones, and vice versa.

In line with the findings regarding the role of chronotype in breast cancer, network-based GWAS studies conducted to identify genetic determinants of breast cancer survival have reported circadian enrichment (63), but only in oestrogen-receptor (ER) negative breast cancers. However, in our previous study, the effect of chronotype on breast cancer risk (OR=0.88, 95% CI:0.82, 0.93) was largely unchanged when stratified into ER-positive (OR= 0.86, 95% CI:0.80, 0.92) or ER-negative breast cancer (OR=0.88, 95% CI:0.80, 0.97)(10). We have extended this to also investigate the effect of chronotype on five subtypes of breast cancer: three luminal-type cancers which are ER-positive, and HER2-enriched and triple negative cancers which are ER-negative. While these results are largely consistent with our findings for a chronotype effect on all breast cancer subtypes (supplementary figure 2), there was some indication for an increased risk of luminal B HER2-negative breast cancer in relation to morning preference, although this was imprecisely estimated.

The effect of both testosterone and SHBG on ER-stratified breast cancer has been previously studied(26), in which both total (OR=1.19, 95% CI:1.12, 1.26) and bioavailable testosterone (OR=1.23, 95% CI:1.12, 1.34) were associated with increased risk of ER-positive breast cancers. This is consistent with our subtype-specific analyses for luminal-type breast cancers (supplementary figure 3A and 3B). In the previous study, there was limited evidence for an association for either total (OR=0.99, 95% CI:0.93, 1.06) or bioavailable testosterone (OR=0.94, 95% CI:0.83, 1.07) on ER-negative breast cancer. While this is consistent with our finding for total testosterone and HER2-enriched breast cancer risk (OR=0.98, 95% CI:0.81, 1.18), there was an indication that bioavailable testosterone reduced risk of HER2-enriched breast cancer (OR=0.86, 95% CI:0.69, 1.07) although this was imprecisely estimated. Similarly, a reduction in risk of triple negative breast cancer was found for both total (OR=0.91, 95% CI:0.81, 1.03) and bioavailable testosterone (OR=0.89, 95% CI:0.77, 1.04). The previous study also reported reduced risk of ER-positive (OR=0.91, 95% CI: 0.82, 1.01) and increased risk of ER-negative (OR=1.14, 95% CI: 1.01, 1.28) breast cancer per SD increase in SHBG. These are consistent with findings from our present study, except for luminal-B HER2-negative breast cancer (ER-positive), for which we found increased risk per SD increase in SHBG, although imprecisely estimated (supplementary figure 3C).

### Strengths & Limitations

A key strength of this study is the use of MR to appraise the causal effect of chronotype on both sex and prostate cancer, as well as additional MR analyses to assess the role of sex hormones underlying these effects. Furthermore, the sensitivity analyses conducted in tandem to the main analyses served to test the assumptions of MR and provide evidence for the robustness of the results. The genetic summary data used for both breast and prostate cancer in this study were obtained from the largest available GWAS available to date and for breast cancer enabled the stratification of breast cancer outcomes into five distinct subtypes, thus improving the clinical relevance of these findings.

While we were careful to use genetic variants from sex-specific GWAS when evaluating the sex-hormones in relation to breast and prostate cancer, the SNPs used to instrument chronotype in the main analyses were extracted from a sex-combined GWAS including data from UKB and 23andMe(32). We used male and female effect estimates for the SNPs derived from UK Biobank in the MR analyses, although the correlation in effect estimates of the SNPs between males and females was only moderate (0.60), suggesting some heterogeneity in the genetic contribution to chronotype between sexes. Given this, we also derived sex-specific instruments and compared results of MR using the combined- and sex-specific chronotype instruments, with the effect estimates obtained being largely consistent with respect to both breast and prostate cancer.

While we conducted a series of sensitivity analyses to assess MR and two-sample MR assumptions, we did not directly test the assumption of independence (IVs must be independent of confounders of the exposure-outcome association). However, germline genetic variants used in MR should not plausibly be influenced by confounding factors, and as such, concerns about violation of the second MR assumption usually focus on confounding by population stratification or as a result of selection bias(64). Steps were taken to minimise population stratification in the GWAS performed although this assumption is difficult to test, particularly in a two-sample setting. Other assumptions for causal effect estimation also exist, such as the causal estimates being homogeneous in the population(65), which we were also not able to explicitly assess.

In terms of two-sample MR specific assumptions, the use of overlapping sample populations between exposures and outcomes may also introduce bias(52). For analyses where exposures were generated from UKB data and outcomes from BCAC or ELLIPSE/PRACTICAL there was no known overlap between participants. However, for bidirectional MR analyses, both exposure and outcomes were from UKB and as such had a 100% overlap. This has been addressed with the use of split-sample MR analysis. While this approach may reduce power, the comparison between whole- and split- sample analyses served as an indicator of the extent to which the main analyses are influenced by this bias. Overall, the results were consistent between these analyses, suggesting that the influence of bias from using overlapping datasets is minimal. Data from UKB may also be influenced by selection bias, however this is a more general issue underlying all UKB data due to low participation rates.

Another potential concern is the overlap in the UKB sample used for both conducting the discovery GWAS for chronotype and sex hormones and for obtaining the effect estimates for the genetic instruments in MR analysis, given the potential for Winner’s curse bias. For chronotype, this was addressed previously(10) whereby sensitivity analyses using only the SNPs that replicated in 23andMe were found to give similar results. However, in the absence of a large independent genetic study with hormone measures, this was not possible for the hormone instruments used in this study.

### Further Work

The analyses presented here demonstrate inconsistent evidence for the role of testosterone as a mediator for the effect of chronotype on cancer, therefore further investigation is required. For example, to improve the robustness of the findings in this study, it would be interesting to investigate the effect of morning-preference on breast cancer subtypes using more objective measures of chronotype, such as L5 timing (66). We believe further work investigating testosterone in relation to other sleep traits (e.g. sleep duration and insomnia) would also be valuable in understanding the nuanced relationship between sleep and cancer risk. Due to the lack of SNPs reaching genome-wide significance, we were unable to sufficiently explore the mediating effect of oestradiol in this study. As such, should a large-scale genetic study of oestradiol data become available, further investigation is warranted. While our study focused on the putative role of sex hormones in the link between chronotype and cancer risk, other hormones such as cortisol and melatonin, have been strongly linked with circadian traits and may lay on the causal pathway to breast cancer. MR analysis using genetic instruments for these hormones is therefore required.

## Conclusion

In conclusion, this study has extended previous findings regarding the protective effect of chronotype on breast cancer and has found evidence to suggest that morning-preference also reduces prostate cancer risk in men. While testosterone levels were found to be closely linked with both chronotype and cancer risk, there was inconsistent evidence for the role of testosterone in mediating the effect of morning preference chronotype on both breast and prostate cancer. Findings regarding the potential protective effect of chronotype on both breast and prostate cancer risk are clinically interesting although this may not serve as a direct target for intervention, since it is difficult to modify someone’s morning/evening preference. Given this, further studies are needed to investigate the mechanisms underlying this effect and to identify potential modifiable intermediates.

## Data Availability

Summary data for the GWAS conducted as part of this study have been made available on the OpenGWAS database (gwas.mrcieu.ac.uk)
This research was conducted using the UK Biobank Resource under application number 16391.
Summary data for the breast cancer GWAS used in this study is available at: http://bcac.ccge.medschl.cam.ac.uk/bcacdata/oncoarray/oncoarray-and-combined-summary-result/gwas-summary-associations-breast-cancer-risk-2020/
Summary data for the prostate cancer GWAS used in this study is available at: http://practical.icr.ac.uk/blog/?page_id=8164

http://gwas.mrcieu.ac.uk

http://bcac.ccge.medschl.cam.ac.uk/bcacdata/oncoarray/oncoarray-and-combined-summary-result/gwas-summary-associations-breast-cancer-risk-2020/

http://practical.icr.ac.uk/blog/?page_id=8164

## Notes

FUNDING BH is funded by an Above & Beyond breast cancer legacy grant from University Hospitals Bristol NHS Foundation Trust. RCR is a de Pass Vice Chancellor’s Research Fellow. RMM is supported by a Cancer Research UK (C18281/A19169) programme grant (the Integrative Cancer Epidemiology Programme). SK is supported by the United Kingdom Research and Innovation Future Leaders Fellowship (MR/T043202/1). KR is funded through the Bristol ICEP2 programme (Cancer Research UK C18281/A29019). BH, TR, SK, RMM, DAL and RCR work in in the Medical Research Council Integrative Epidemiology Unit at the University of Bristol supported by the Medical Research Council (MC_UU_00011/1 and MC_UU_00011/6) and the University of Bristol. RMM and DAL also supported by the National Institute for Health Research (NIHR) Bristol Biomedical Research Centre, which is funded by the National Institute for Health Research (NIHR) and is a partnership between University Hospitals Bristol NHS Foundation Trust and the University of Bristol.

CONFLICT OF INTEREST DAL has received support from Roche Diagnostics and Medtronic Ltd for research unrelated to that presented here. TR has received grants from Daiichi-Sankyo and Amgen to attend educational workshops. All other authors have no competing interests to declare. The funders had no role in the design of the study; the collection, analysis, and interpretation of the data; the writing of the manuscript; and the decision to submit the manuscript for publication. The authors alone are responsible for the views expressed in this article.

### Competing Interest Statement

DAL has received support from Roche Diagnostics and Medtronic Ltd for research unrelated to that presented here. TR has received grants from Daiichi-Sankyo and Amgen to attend educational workshops. All other authors have no competing interests to declare. The funders had no role in the design of the study; the collection, analysis, and interpretation of the data; the writing of the manuscript; and the decision to submit the manuscript for publication. The authors alone are responsible for the views expressed in this article.

### Funding Statement

BH is funded by an Above & Beyond breast cancer legacy grant from University Hospitals Bristol NHS Foundation Trust. RCR is a de Pass Vice Chancellor's Research Fellow. RMM is supported by a Cancer Research UK (C18281/A19169) programme grant (the Integrative Cancer Epidemiology Programme). SK is supported by the United Kingdom Research and Innovation Future Leaders Fellowship (MR/T043202/1). KR is funded through the Bristol ICEP2 programme (Cancer Research UK C18281/A29019). BH, TR, SK, RMM, DAL and RCR work in in the Medical Research Council Integrative Epidemiology Unit at the University of Bristol supported by the Medical Research Council (MC_UU_00011/1 and MC_UU_00011/6) and the University of Bristol. RMM and DAL also supported by the National Institute for Health Research (NIHR) Bristol Biomedical Research Centre, which is funded by the National Institute for Health Research (NIHR) and is a partnership between University Hospitals Bristol NHS Foundation Trust and the University of Bristol.

### Author Declarations

The study was approved by the UKB data access committee, and informed consent was obtained from all participants. All participating ELLIPSE, PRACTICAL and BCAC studies were approved by their appropriate ethics or institutional review board and all participants provided informed consent.

## References

1. Ferlay J, Colombet M, Soerjomataram I, Mathers C, Parkin DM, Piñeros M, et al. Estimating the global cancer incidence and mortality in 2018: GLOBOCAN sources and methods. Int J Cancer. 2019;144(8):1941–53.

2. American Cancer Society. About Breast Cancer. Breast Cancer Facts Fig. 2017;1–19.

3. Nuhn P, De Bono JS, Fizazi K, Freedland SJ, Grilli M, Kantoff PW, et al. Update on Systemic Prostate Cancer Therapies: Management of Metastatic Castration-resistant Prostate Cancer in the Era of Precision Oncology. Eur Urol. 2019;75(1):88–99.

4. Piotrzkowska-Wróblewska H, Dobruch-Sobczak K, Klimonda Z, Karwat P, Roszkowska-Purska K, Gumowska M, et al. Monitoring breast cancer response to neoadjuvant chemotherapy with ultrasound signal statistics and integrated backscatter. PLoS One. 2019 Mar 14;14(3):e0213749.

5. Gourd E. New advances in prostate cancer screening and monitoring. Lancet Oncol. 2020;21(7):887.

6. Smittenaar CR, Petersen KA, Stewart K, Moitt N. Cancer incidence and mortality projections in the UK until 2035. Br J Cancer. 2016/10/11. 2016 Oct 25;115(9):1147–55.

7. He C, Anand ST, Ebell MH, Vena JE, Robb SW. Circadian disrupting exposures and breast cancer risk: a meta-analysis. Int Arch Occup Environ Health. 2015;88(5):533–47.

8. Sigurdardottir LG, Valdimarsdottir UA, Fall K, Rider JR, Lockley SW, Schernhammer E, et al. Circadian disruption, sleep loss, and prostate cancer risk: A systematic review of epidemiologic studies. Cancer Epidemiol Biomarkers Prev. 2012;21(7):1002–11.

9. Adan A, Natale V. Gender differences in morningness-eveningness preference. Chronobiol Int. 2002;19(4):709–20.

10. Richmond RC, Anderson EL, Dashti HS, Jones SE, Lane JM, Strand LB, et al. Investigating causal relations between sleep traits and risk of breast cancer in women: Mendelian randomisation study. BMJ. 2019;365:1–12.

11. Papantoniou K, Castaño-Vinyals G, Espinosa A, Aragonés N, Pérez-Gõmez B, Burgos J, et al. Night shift work, chronotype and prostate cancer risk in the MCC-Spain case-control study. Int J Cancer. 2015;137(5):1147–57.

12. Kalmbach DA, Schneider LD, Cheung J, Bertrand SJ, Kariharan T, Pack AI, et al. Genetic Basis of Chronotype in Humans: Insights From Three Landmark GWAS. Sleep. 2017 Feb 1;40(2):zsw048.

13. Shawa N, Rae DE, Roden LC. Impact of seasons on an individual’s chronotype: current perspectives. Nat Sci Sleep. 2018 Oct 31;10:345–54.

14. Randler C, Faßl C, Kalb N. From Lark to Owl: Developmental changes in morningness-eveningness from new-borns to early adulthood. Sci Rep. 2017;7(January):1–8.

15. Roenneberg T, Kuehnle T, Pramstaller PP, Ricken J, Havel M, Guth A, et al. A marker for the end of adolescence. Curr Biol. 2004;14(24):R1038–9.

16. Randler C, Schaal S. Morningness-eveningness, habitual sleep-wake variables and cortisol level. Biol Psychol. 2010 Sep;85(1):14–8.

17. Premkumar M, Sable T, Dhanwal D, Dewan R. Circadian Levels of Serum Melatonin and Cortisol in relation to Changes in Mood, Sleep, and Neurocognitive Performance, Spanning a Year of Residence in Antarctica. Neurosci J. 2012/12/13. 2013;2013:254090.

18. Maierova L, Borisuit A, Scartezzini J-L, Jaeggi SM, Schmidt C, Münch M. Diurnal variations of hormonal secretion, alertness and cognition in extreme chronotypes under different lighting conditions. Sci Rep. 2016 Sep 20;6:33591.

19. Randler C, Ebenhöh N, Fischer A, Höchel S, Schroff C, Stoll JC, et al. Chronotype but not sleep length is related to salivary testosterone in young adult men. Psychoneuroendocrinology. 2012;37(10):1740–4.

20. Jankowski KS, Fajkowska M, Domaradzka E, Wytykowska A. Chronotype, social jetlag and sleep loss in relation to sex steroids. Psychoneuroendocrinology. 2019;108(May):87–93.

21. Davey Smith G, Ebrahim S. ‘Mendelian randomization’: can genetic epidemiology contribute to understanding environmental determinants of disease?*. Int J Epidemiol. 2003 Feb 1;32(1):1–22.

22. Davey Smith G, Hemani G. Mendelian randomization: genetic anchors for causal inference in epidemiological studies. Hum Mol Genet. 2014/07/04. 2014 Sep 15;23(R1):R89–98.

23. Davies NM, Holmes M V., Davey Smith G. Reading Mendelian randomisation studies: A guide, glossary, and checklist for clinicians. BMJ. 2018;362.

24. Pierce BL, Burgess S. Efficient design for Mendelian randomization studies: subsample and 2-sample instrumental variable estimators. Am J Epidemiol. 2013 Oct;178(7):1177–84.

25. Dimou NL, Papadimitriou N, Gill D, Christakoudi S, Murphy N, Gunter MJ, et al. Sex hormone binding globulin and risk of breast cancer: a Mendelian randomization study. Int J Epidemiol. 2019 Jun 1;48(3):807–16.

26. Ruth KS, Day FR, Tyrrell J, Thompson DJ, Wood AR, Mahajan A, et al. Using human genetics to understand the disease impacts of testosterone in men and women. Nat Med. 2020;26(2):252–8.

27. Collaborative Group on Hormonal Factors in Breast Cancer. Type and timing of menopausal hormone therapy and breast cancer risk: individual participant meta-analysis of the worldwide epidemiological evidence. Lancet. 2019;394(10204):1159–68.

28. Key TJ, Appleby PN, Reeves GK, Travis RC, Brinton LA, Helzlsouer KJ, et al. Steroid hormone measurements from different types of assays in relation to body mass index and breast cancer risk in postmenopausal women: Reanalysis of eighteen prospective studies. Steroids. 2014/10/07. 2015 Jul;99(Pt A):49–55.

29. Zhang H, Ahearn TU, Lecarpentier J, Barnes D, Beesley J, Qi G, et al. Genome-wide association study identifies 32 novel breast cancer susceptibility loci from overall and subtype-specific analyses. Nat Genet. 2020;52(6):572–81.

30. Burgess S, Thompson SG. Multivariable Mendelian randomization: the use of pleiotropic genetic variants to estimate causal effects. Am J Epidemiol. 2015/01/27. 2015 Feb 15;181(4):251–60.

31. Sanderson E, Davey Smith G, Windmeijer F, Bowden J. An examination of multivariable Mendelian randomization in the single-sample and two-sample summary data settings. Int J Epidemiol. 2019 Jun 1;48(3):713–27.

32. Jones SE, Lane JM, Wood AR, van Hees VT, Tyrrell J, Beaumont RN, et al. Genome-wide association analyses of chronotype in 697,828 individuals provides insights into circadian rhythms. Nat Commun. 2019;10(1).

33. Loh PR, Tucker G, Bulik-Sullivan BK, Vilhjálmsson BJ, Finucane HK, Salem RM, et al. Efficient Bayesian mixed-model analysis increases association power in large cohorts. Nat Genet. 2015;47(3):284–90.

34. Michailidou K, Lindström S, Dennis J, Beesley J, Hui S, Kar S, et al. Association analysis identifies 65 new breast cancer risk loci. Nature. 2017;551(7678):92–4.

35. Al Olama AA, Kote-Jarai Z, Berndt SI, Conti D V, Schumacher F, Han Y, et al. A meta-analysis of 87,040 individuals identifies 23 new susceptibility loci for prostate cancer. Nat Genet. 2014/09/14. 2014 Oct;46(10):1103–9.

36. Schumacher FR, Al Olama AA, Berndt SI, Benlloch S, Ahmed M, Saunders EJ, et al. Association analyses of more than 140,000 men identify 63 new prostate cancer susceptibility loci. Nat Genet. 2018 Jul;50(7):928–36.

37. Jones SE, Lane JM, Wood AR, van Hees VT, Tyrrell J, Beaumont RN, et al. Genome-wide association analyses of chronotype in 697,828 individuals provides insights into circadian rhythms. Nat Commun. 2019;10(1):343.

38. UK Biobank. Biomarker assay quality procedures: approaches used to minimise systematic and random errors (and the wider epidemiological implications). 2019;(April):25.

39. Wald A. The Fitting of Straight Lines if Both Variables are Subject to Error. Ann Math Stat. 1940 Jan 22;11(3):284–300.

40. Davies NM, Holmes M V., Davey Smith G. Reading Mendelian randomisation studies: A guide, glossary, and checklist for clinicians. BMJ. 2018;362.

41. Burgess S, Thompson SG. Avoiding bias from weak instruments in mendelian randomization studies. Int J Epidemiol. 2011;40(3):755–64.

42. Society TE. Instrumental Variables Regression with Weak Instruments Author (s): Douglas Staiger and James H. Stock Reviewed work (s): Published by?: The Econometric Society Stable URL?: http://www.jstor.org/stable/2171753. Society. 2011;65(3):p557–86.

43. Sanderson E, Spiller W, Bowden J. Testing and Correcting for Weak and Pleiotropic Instruments in Two-Sample Multivariable Mendelian Randomisation. bioRxiv. 2020 Jan 1;2020.04.02.021980.

44. Bowden J, Davey Smith G, Burgess S. Mendelian randomization with invalid instruments: effect estimation and bias detection through Egger regression. Int J Epidemiol. 2015 Apr;44(2):512–25.

45. Bowden J, Davey Smith G, Haycock PC, Burgess S. Consistent Estimation in Mendelian Randomization with Some Invalid Instruments Using a Weighted Median Estimator. Genet Epidemiol. 2016 May;40(4):304–14.

46. Hartwig FP, Davey Smith G, Bowden J. Robust inference in summary data Mendelian randomization via the zero modal pleiotropy assumption. Int J Epidemiol. 2017 Dec 1;46(6):1985–98.

47. Cochran WG. The Combination of Estimates from Different Experiments. Biometrics. 1954 Sep 29;10(1):101–29.

48. Greco M F Del, Minelli C, Sheehan NA, Thompson JR. Detecting pleiotropy in Mendelian randomisation studies with summary data and a continuous outcome. Stat Med. 2015;34(21):2926–40.

49. Bowden J, Spiller W, Del Greco M F, Sheehan N, Thompson J, Minelli C, et al. Improving the visualization, interpretation and analysis of two-sample summary data Mendelian randomization via the Radial plot and Radial regression. Int J Epidemiol. 2018 Aug;47(4):1264–78.

50. Blakely T, McKenzie S, Carter K. Misclassification of the mediator matters when estimating indirect effects. J Epidemiol Community Health. 2013 May;67(5):458–66.

51. Hemani G, Tilling K, Davey Smith G. Orienting the causal relationship between imprecisely measured traits using GWAS summary data. PLOS Genet. 2017 Nov 17;13(11):e1007081.

52. Burgess S, Davies NM, Thompson SG. Bias due to participant overlap in two-sample Mendelian randomization. Genet Epidemiol. 2016/09/14. 2016 Nov;40(7):597–608.

53. Higgins JPT, Thompson SG. Quantifying heterogeneity in a meta-analysis. Stat Med. 2002 Jun 15;21(11):1539–58.

54. Miller MA, Kruisbrink M, Wallace J, Ji C, Cappuccio FP. Sleep duration and incidence of obesity in infants, children, and adolescents: a systematic review and meta-analysis of prospective studies. Sleep. 2018 Apr;41(4).

55. Wang J, Li AM, Lam HSHS, Leung GM, Schooling CM. Sleep Duration and Adiposity in Children and Adults: Observational and Mendelian Randomization Studies. Obesity. 2019 Jun 1;27(6):1013–22.

56. Stanikova D, Luck T, Pabst A, Bae YJ, Hinz A, Glaesmer H, et al. Associations Between Anxiety, Body Mass Index, and Sex Hormones in Women. Vol. 10, Frontiers in Psychiatry. 2019. p. 479.

57. Shamim MO, Ali Khan FM, Arshad R. Association between serum total testosterone and Body Mass Index in middle aged healthy men. Pakistan J Med Sci. 2015;31(2):355–9.

58. Akin F, Bastemir M, Alkis E, Kaptanoglu B. Associations between sex hormone binding globulin and metabolic syndrome parameters in premenopausal obese women. Indian J Med Sci. 2008 Oct;62(10):407–15.

59. Amin HA, Kaewsri P, Yiorkas AM, Cooke H, Blakemore AI, Drenos F. Increased adiposity is protective for breast and prostate cancer: a Mendelian randomisation study using up to 132,413 breast cancer cases and 85,907 prostate cancer cases. medRxiv. 2020 Jan 1;2020.04.02.20049031.

60. Hemani G, Zheng J, Elsworth B, Wade KH, Haberland V, Baird D, et al. The MR-Base platform supports systematic causal inference across the human phenome. Elife. 2018 May;7.

61. Locke AE, Kahali B, Berndt SI, Justice AE, Pers TH, Day FR, et al. Genetic studies of body mass index yield new insights for obesity biology. Nature. 2015 Feb 12;518(7538):197–206.

62. Sanderson E, Davey Smith G, Windmeijer F, Bowden J. An examination of multivariable Mendelian randomization in the single-sample and two-sample summary data settings. Int J Epidemiol. 2018 Dec 10;48(3):713–27.

63. Escala-Garcia M, Abraham J, Andrulis IL, Anton-Culver H, Arndt V, Ashworth A, et al. A network analysis to identify mediators of germline-driven differences in breast cancer prognosis. Nat Commun. 2020;11(1):1–14.

64. Yang Q, Sanderson E, Tilling K, Borges MC, Lawlor DA. Exploring and mitigating potential bias when genetic instrumental variables are associated with multiple non-exposure traits in Mendelian randomization. medRxiv. 2019 Jan 1;19009605.

65. Labrecque J, Swanson SA. Understanding the Assumptions Underlying Instrumental Variable Analyses: a Brief Review of Falsification Strategies and Related Tools. Curr Epidemiol reports. 2018;5(3):214–20.

66. Jones SE, van Hees VT, Mazzotti DR, Marques-Vidal P, Sabia S, van der Spek A, et al. Genetic studies of accelerometer-based sleep measures yield new insights into human sleep behaviour. Nat Commun. 2019;10(1):1–12.

