## Supplementary Figures for "Do sex hormones confound or mediate the effect of chronotype on breast and prostate cancer? A Mendelian randomization study"

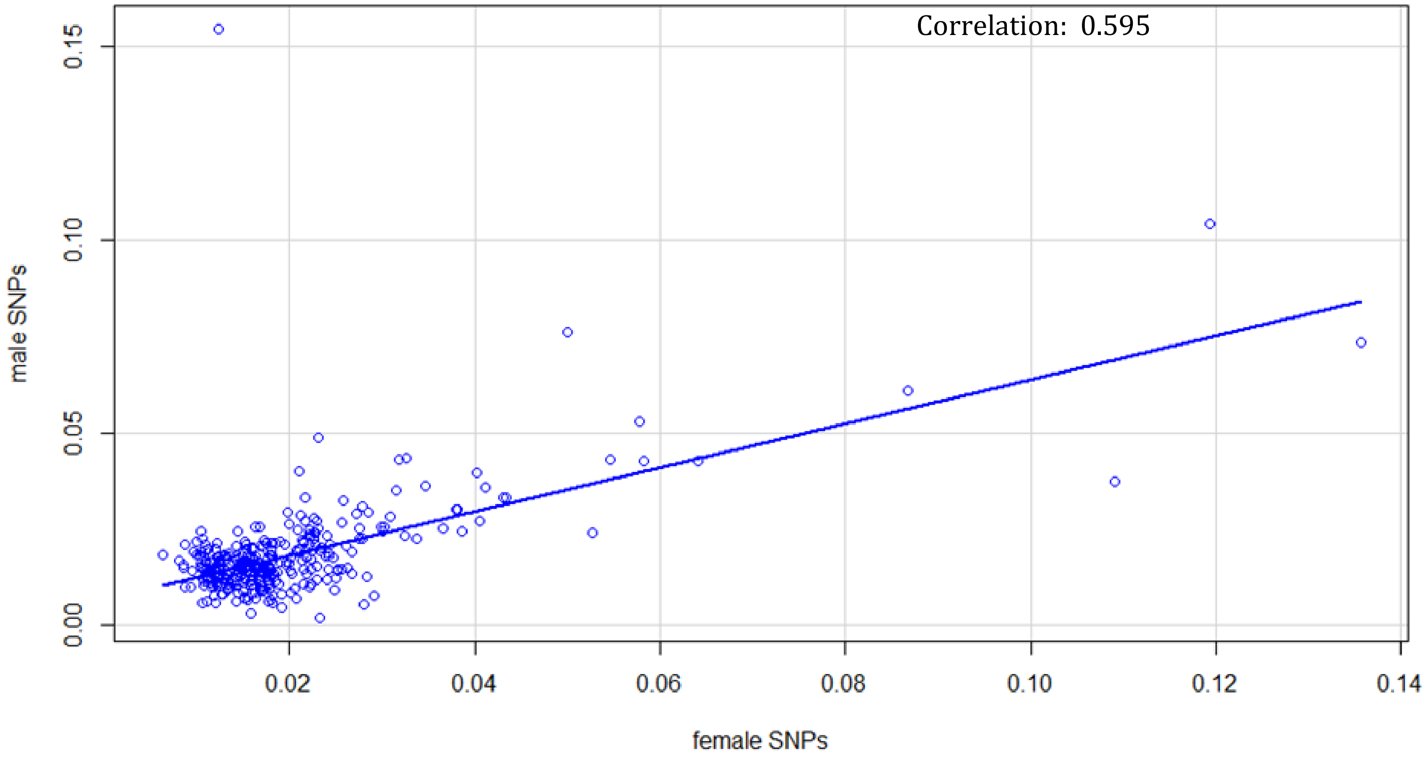

Supplementary figure 1. Scatter plot of genetic correlation between male and female effect estimates for chronotype instrument.

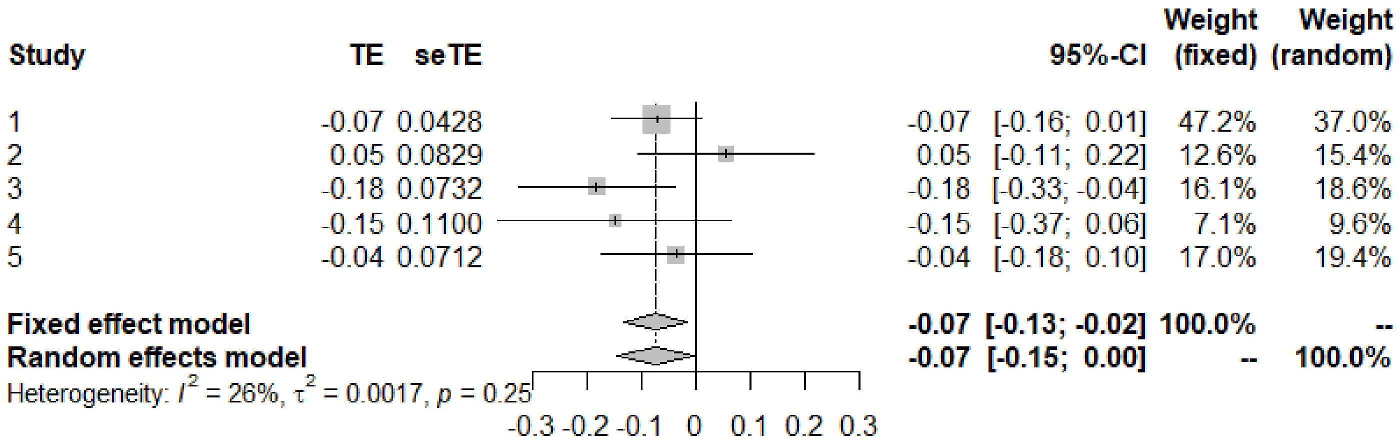


Supplementary figure 2. Heterogeneity analysis for MR of chronotype on breast cancer subtypes. Study numbers = 1. Luminal A (ER+/PR+, HER2-); 2. Luminal B (ER+/PR+/-, HER2+); 3. Luminal B (ER+/PR+/-, HER2-, Ki 67 > 14%); 4. HER2 (ER-, PR-, HER2+); and 5. Triple Negative (ER-, PR-, HER2-).

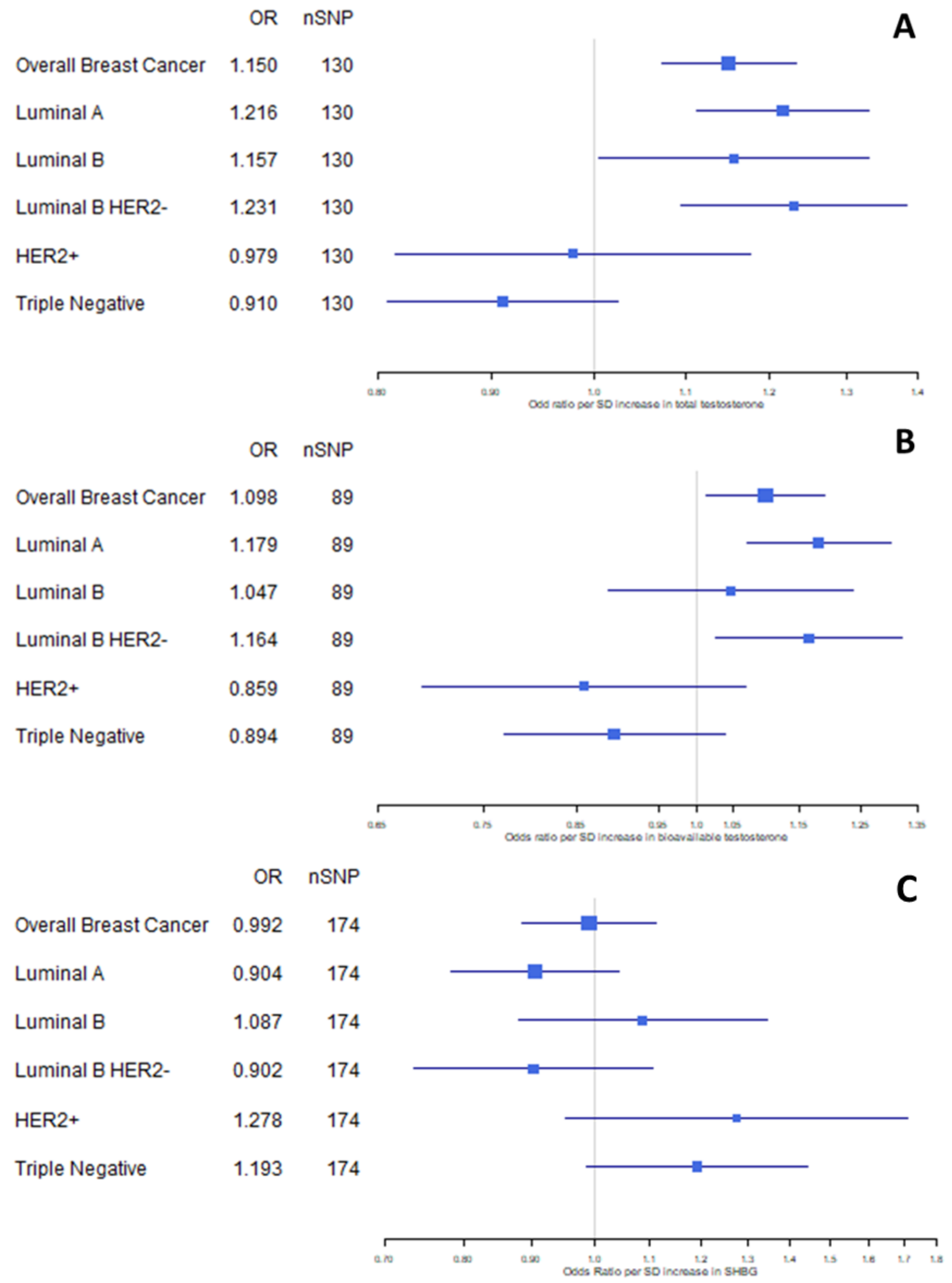


Supplementay figure 3. Forest plot of uvMR results for A) total testosterone, B) bioavailable testosterone and C) SHBG effects on breast cancer subtypes.

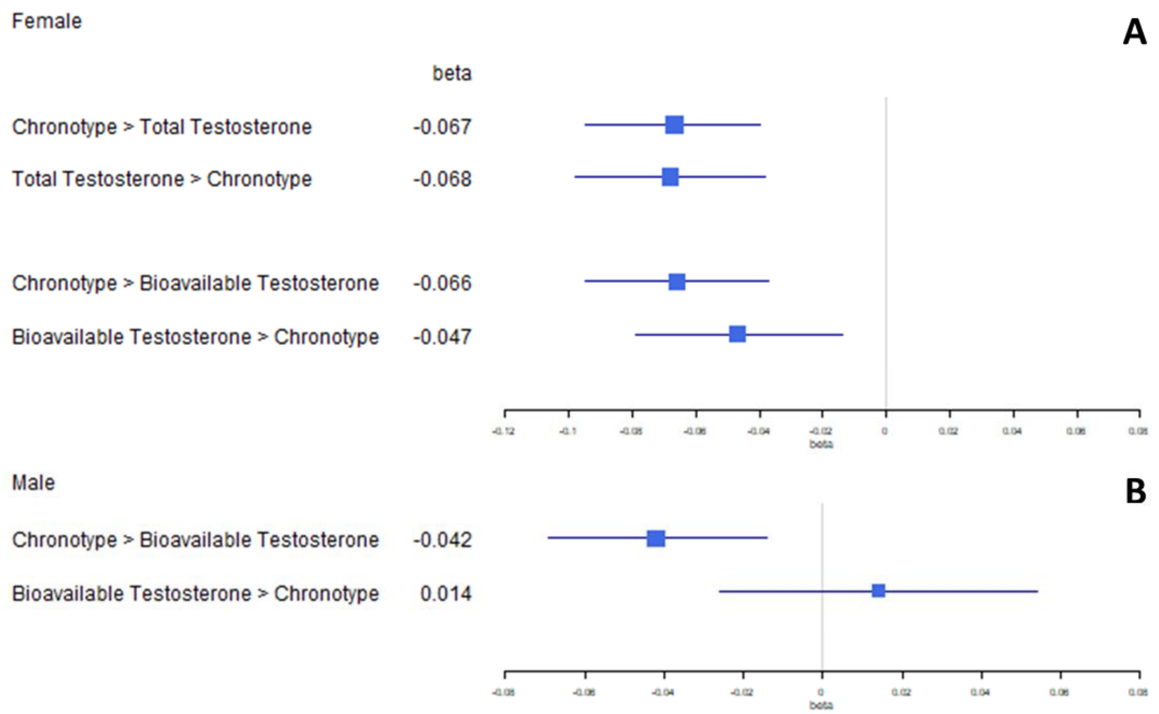


Supplementary figure 4. Forest plot of bdMR results for chronotype and sex hormones in female (A) and male (B) datasets. All instruments in these analyses have been subjected to Steiger-filtering.

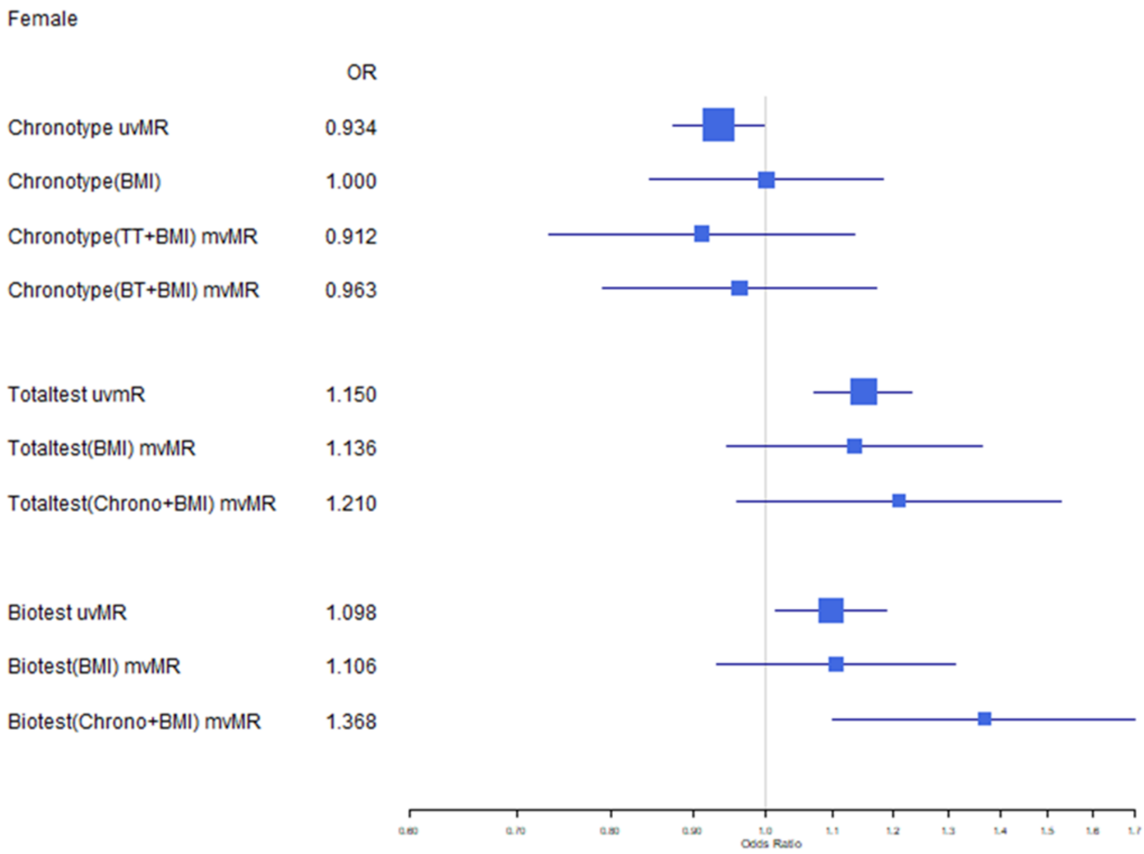


Supplementary figure 5. Forest plot of bivariate (adjusted for BMI only) and trivariate (adjusted for chronotype/sex hormone and BMI) mvMR results in females.

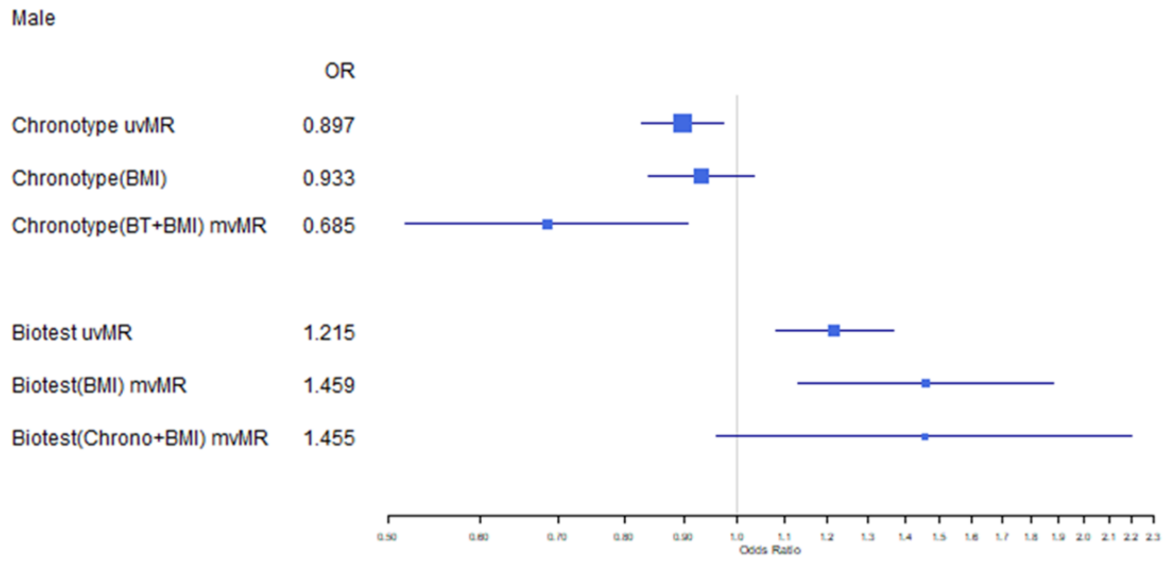


Supplementary figure 6. Forest plot of bivariate (adjusted for BMI only) and trivariate (adjusted for chronotype/sex hormone and BMI) mvMR results in males.
