## Supplementary Methods for "Do sex hormones confound or mediate the effect of chronotype on breast and prostate cancer? A Mendelian randomization study"

Univariable two-sample MR analysis was performed to estimate the effects of chronotype, total testosterone, bioavailable testosterone, SHBG and oestradiol on breast and prostate cancer using SNP-exposure estimates from UK Biobank (sample 1) and SNP-outcome estimates (sample 2) extracted from summary statistics obtained from the Breast Cancer Association Consortium (BCAC)[1,2], Elucidating Loci Involved in Prostate Cancer (ELLIPSE) and The Prostate Cancer Association Group to Investigate Cancer Associated Alterations in the Genome (PRACTICAL) consortia[3,4] (PRACTICAL/ELLIPSE) GWAS studies.

Univariable MR analyses were also conducted using data stratified by breast cancer subtype[2] (luminal A, luminal B, luminal B HER2 negative, HER2-enriched and triple negative).

All univariable MR analyses used the R package "Two SampleMR"[5]. If SNPs within the exposure instrument were not present in the outcome (breast and prostate cancer) GWAS, proxy SNPs with a minimum linkage disequilibrium (LD) of $r2 = 0.8$ were substituted. Exposure and outcome SNPs were harmonised to ensure effect estimates were aligned with the same reference allele for both datasets. This was inferred using allele frequencies for palindromes, and if direction of effect could not be inferred (MAF>0.42), the SNP was excluded. The mr() function was then used to obtain a random-effects inverse variance weighted (IVW) estimate for the causal effect of the exposure on the outcome.

Bidirectional MR: Reciprocal Effects between Chronotype and Sex Hormones

In bidirectional MR analyses, we performed MR to evaluate the reciprocal effect of each hormone on chronotype. Together with the effect obtained from the MR of chronotype on hormones, these estimates can be used to determine directionality of the relationship. To ensure the robustness of the inferred direction between chronotype and the sex hormone traits, the instruments were subjected to Steiger filtering to assess strength of SNP associations with both exposure and outcome[6]. Any SNPs found to be more strongly associated with the outcome than the exposure were subsequently removed from the instrument, to ensure that the instrument is only influencing the outcome through the exposure of interest. This Steiger filtering of instruments was also subjected to sensitivity testing (see 'sensitivity analyses'). The same approach was taken as in the univariable, with effect estimates obtained from IVW analysis.

Multivariable MR: Direct Effects of Chronotype and Sex Hormones on Cancer Risk

Multivariable MR (mvMR) analysis was conducted to assess the direct effects of chronotype and sex hormones on both breast and prostate cancer[7,8]. For these analyses, the instruments for each exposure included in the previous uvMR analyses were used. These instruments were first clumped individually (r2 = 0.001), before being combined and then clumped again[9]. The mv_multiple() function was then used to obtain an inverse variance weighted (IVW) estimate for the direct causal effect of each exposure on the outcome, in which instruments for each exposure are regressed against the outcome together, weighting for the inverse variance of the outcome[5]. Evidence of attenuation of the chronotype effect in mvMR compared to the corresponding uvMR results can be used to indicate a mediated (or indirect) effect executed via the sex hormones investigated in this study[10].

**Supplementary Methods 2: Genotyping, Imputation and association analysis (UKB)**

The full data release contains the cohort of successfully genotyped samples (n=488,377). 49,979 individuals were genotyped using the UK BiLEVE array and 438,398 using the UK Biobank axiom array. Pre-imputation quality control, phasing and imputation are described elsewhere[11]. In brief, prior to phasing, multiallelic SNPs or those with minor allele frequency (MAF) ≤1% were removed. Phasing of genotype data was performed using a modified version of the SHAPEIT2 algorithm[12]. Genotype imputation to a reference set combining the UK10K haplotype and HRC reference panels 8 was performed using IMPUTE2 algorithms[13]. The analyses presented here were restricted to autosomal variants within the HRC site list using a graded filtering with varying imputation quality for different allele frequency ranges. Therefore, rarer genetic variants are required to have a higher imputation INFO score (Info>0.3 for MAF >3%; Info>0.6 for MAF 1-3%; Info>0.8 for MAF 0.5-1%; Info>0.9 for MAF 0.1-0.5%) with MAF and Info scores having been recalculated on an in-house derived 'European' subset[14].

Individuals with sex-mismatch (derived by comparing genetic sex and reported sex) or individuals with sex-chromosome aneuploidy were excluded from the analysis (n=814). We restricted the sample to individuals of 'European' ancestry as defined by an in-house k-means cluster analysis performed using the first 4 principal components provided by UKB in the statistical software environment R (n=464,708)[14].

Genome-wide association analysis (GWAS) was conducted using linear mixed model (LMM) association method as implemented in BOLT-LMM (v2.3)[15]. To model population structure in the sample we used 143,006 directly genotyped SNPs, obtained after filtering on MAF > 0.01; genotyping

UOB Open

rate > 0.015; Hardy-Weinberg equilibrium p-value < 0.0001 and LD pruning to an r2 threshold of 0.1 using PLINKv2.00. Genotype array was adjusted for in the model.

**Supplementary Methods 3: Genotyping and Imputation (BCAC)**

Summary GWAS data obtained from BCAC came from a meta-analysis of 133,384 breast cancer cases and 113,789 controls[1]. Summary statistics were also obtained from analyses of 106,491 cases and 94,407 controls for five breast cancer subtypes (Luminal A (ER+/ PR+, HER2); Luminal B (ER+/PR+/-, HER2+); Luminal B HER2 negative (ER+/PR+/-, HER2-, Ki-67 > 14%); HER2 (ER-, PR-, HER2+); and Triple Negative (ER-, PR-, HER2-))[2]. A subset of samples from the overall breast cancer GWAS were obtained by genotyping with an Illumina custom infinium array (OncoArray) of ~530,000 SNPs. Criteria for removal of SNPs include: i) concordance of <98% among 5,280 duplicate sample pairs; ii) a $p < 1.0 \times 10^{-12}$ in cases or $p < 1.0 \times 10^{-7}$ in controls and; iii) a call rate of <95%. SNPs were also removed where the cluster plot was judged to be inadequate. Samples from the overall breast cancer GWAS were also obtained through imputation using the 1000 genomes project phase 3 reference panel[16], where criteria for removal include: i) minor allele frequency of <1%; ii) a call rate of <98% or; iii) different frequency or absence from the 1000 genomes reference panel.

Results from the most recent OncoArray analyses were combined with 46,785 cases and 42,892 controls from the previous iCOGS genotyping project, plus 46,785 cases and 42,892 controls from eleven other breast cancer GWAS using a fixed-effects meta-analysis. All contributory GWAS were adjusted for principal components of European ancestry. OncoArray analyses were further adjusted for country and iCOGS analyses for study. All participating studies had the approval of their appropriate ethics review board and all participants provided informed consent.

**Supplementary Methods 4: Genotyping and Imputation (ELLIPSE/PRACTICAL)**

The ELLIPSE Consortium includes the meta-analysis of both existing and novel GWAS, plsu iCOGS genotyping. The meta-analysis includes only participants of European ancestry, combining GWAS from: UK GWAS stage 1 (Illumina Infinium HumanHap 550 Array: 1,854 cases and 1,894 controls), UK GWAS stage 2 (Illumina iSELECT: 3,706 cases and 3,884 controls), CAPS1 (Affymetrix GeneChip 500K: 474 cases and 482 controls), CAPS2 (Affymetrix GeneChip 5.0K: 1,458 cases and 512 controls), BPC3 (Illumina Human610 Illumina: 2,068 cases and 3,011 controls), PEGASUS (HumanOmni2.5: 4,600 cases and 2,941 controls). From UKGPCS, genotyping was conducted for 977 prostate cancer cases and exome SNP array genotyping was conducted for 4741 subjects. ICOGs genotyping was conducted for 10,366 subjects (including 1,648 participants from the Multiethnic Cohort and 8,718 from UKGPCS). Duplicate samples and first-degree relatives were excluded both within each study and across GWAS studies. Ancestry was assessed through principal component analysis, and only individuals with an estimated proportion of European ancestry > 0.8 (with reference to HapMap populations) were retained.

OncoArray genotyping of 533,631 selected variants from ELLIPSE studies was conducted across five sites (Cambridge, CIDR, Copenhagen, USC, and NCI).Exclusion criteria for SNPs included: a <95% call rate by study; a <98% call rate in any study; a $p < 1.0 \times 10^{-12}$ in cases or $p < 1.0 \times 10^{-7}$ in controls; concordance of <98% among 11,260 duplicate pairs; a minor allele frequency of <1%; different frequency or absence from the 1000 genomes reference panel or; where the cluster plot was judged to be inadequate. Of the 533,631 SNPs on the OncoArray, 498,417 were retained following quality control. Approximately 70 million SNPs were imputed for all samples using the 1000 genomes project phase 3 reference panel. OncoArray and GWAS datasets were imputed through a two-stage approach, using SHAPEIT31 for phasing and IMPUTEv2[17] for imputation.

**Supplementary Methods 5: Assessment of MR Assumptions**

MR analysis relies on three key assumptions: i) IVs must be robustly associated with the exposure of interest; ii) IVs must be independent of confounders of the exposure-outcome association; iii) IVs must only influence the outcome through the exposure of interest[18].

To test the first of the MR assumptions, F-statistics were calculated for all IVs used in univariable analysis to assess instrument strength. The F-statistic incorporates phenotypic variance explained by genetic variants (r2), sample size, and number of variants to estimate strength of the relationship between IVs and phenotype[19,20]. Instrument strength was also assessed in mvMR analyses with conditional F-statistics[21]. In two-sample MR weak instrument bias is expected to result in attenuation of effects towards the null.

While germline genetic variants should not plausibly be influenced by confounding factors, concerns about violation of the second MR assumption usually focus on confounding by ancestry. Steps were taken to minimise population stratification in the GWAS performed although this assumption is difficult to test, particularly in a two-sample setting.

The third assumption relates to potential horizontal pleiotropy of the IVs used in the genetic variants. In particular, the main IVW approach used assumes that the genetic IVs are valid and that there is no unbalanced horizontal pleiotropy. We undertook complementary sensitivity analyses using methods that relax this assumption. MR-Egger, is similar to IVW but does not constrain the intercept to be zero. A non-zero intercept indicates the presence of horizontal pleiotropy and the slope (effect estimate) has controlled for potential bias due to unbalanced horizontal pleiotropy.[22] Both IVW and MR-Egger assume that the 'instrument strength independent of direct effect' (INSIDE) assumption is not violated (i.e. that there is no correlation between the genetic IV-exposure association and unmediated IV-outcome association). The weighted median method assumes that ≥50% of the weight in the analysis stems from IVs which are valid (i.e. if one strong instrument reflecting 50% or more of the variation in the exposure is pleiotropic or several weaker instruments together explain 50% or more of the association and are pleiotropic, the estimate will be biased)[23]. Weighted mode-based MR, estimates from individual valid IVs are in the majority[24]. Scatter plots were used to visualise consistency between IVW, MR-Egger, median and mode effect estimates for initial uvMR of chronotype and sex hormone measures on breast and prostate cancer risk. In subsequent bdMR, and mvMR, MR-Egger alone was used to estimates effects which were robust to pleiotropy.

Steiger filtering was applied to instruments in the bdMR analysis of chronotype and sex hormones. This approach removes SNPs that explain more of the variance in the outcome than the exposure and consequently reduces the likelihood of erroneous results due to pleiotropy [6]. Steiger sensitivity ratios (R) were also calculated for these analyses, which considers the measurement precision of both exposure and outcome, and represents the likelihood that the observed direction of association is correct[25]. An R>1 provides evidence to support the direction of association from exposure to outcome.

1. Michailidou, K. *et al.* Association analysis identifies 65 new breast cancer risk loci. *Nature* **551**, 92–94 (2017).

2. Zhang, H. *et al.* Genome-wide association study identifies 32 novel breast cancer susceptibility loci from overall and subtype-specific analyses. *Nat. Genet.* **52**, 572–581 (2020).

3. Al Olama, A. A. *et al.* A meta-analysis of 87,040 individuals identifies 23 new susceptibility loci for prostate cancer. *Nat. Genet.* **46**, 1103–1109 (2014).

4. Schumacher, F. R. *et al.* Association analyses of more than 140,000 men identify 63 new prostate cancer susceptibility loci. *Nat. Genet.* **50**, 928–936 (2018).

5. Hemani, G. *et al.* The MR-Base platform supports systematic causal inference across the human phenome. *Elife* **7**, (2018).

6. Blakely, T., McKenzie, S. & Carter, K. Misclassification of the mediator matters when estimating indirect effects. *J. Epidemiol. Community Health* **67**, 458–466 (2013).

7. Burgess, S., Daniel, R. M., Butterworth, A. S. & Thompson, S. G. Network Mendelian randomization: using genetic variants as instrumental variables to investigate mediation in causal pathways. *Int. J. Epidemiol.* **44**, 484–495 (2015).

8. Sanderson, E., Davey Smith, G., Windmeijer, F. & Bowden, J. An examination of multivariable Mendelian randomization in the single-sample and two-sample summary data settings. *Int. J. Epidemiol.* **48**, 713–727 (2019).

9. Sanderson, E., Davey Smith, G., Windmeijer, F. & Bowden, J. An examination of multivariable Mendelian randomization in the single-sample and two-sample summary data settings. *Int. J. Epidemiol.* **48**, 713–727 (2018).

10. VanderWeele, T. J. Mediation Analysis: A Practitioner's Guide. *Annu. Rev. Public Health* **37**, 17–32 (2016).

11. Bycroft, C. *et al.* The UK Biobank resource with deep phenotyping and genomic data. *Nature* **562**, 203–209 (2018).

12. O'Connell, J. *et al.* Haplotype estimation for biobank-scale data sets. *Nat. Genet.* **48**, 817–820 (2016).

13. Howie, B., Marchini, J. & Stephens, M. Genotype imputation with thousands of genomes. *G3 (Bethesda).* **1**, 457–470 (2011).

14. Mitchell, R. *et al.* MRC IEU UK Biobank GWAS pipeline version 2. University of Bristol. *Univ. Bristol* (2019).

15. Loh, P. R. *et al.* Efficient Bayesian mixed-model analysis increases association power in large cohorts. *Nat. Genet.* **47**, 284–290 (2015).

UOB Open

16. Auton, A. *et al.* A global reference for human genetic variation. *Nature* **526**, 68–74 (2015).

17. Howie, B. N., Donnelly, P. & Marchini, J. A flexible and accurate genotype imputation method for the next generation of genome-wide association studies. *PLoS Genet.* **5**, e1000529 (2009).

18. Davies, N. M., Holmes, M. V. & Davey Smith, G. Reading Mendelian randomisation studies: A guide, glossary, and checklist for clinicians. *BMJ* **362**, (2018).

19. Burgess, S. & Thompson, S. G. Avoiding bias from weak instruments in mendelian randomization studies. *Int. J. Epidemiol.* **40**, 755–764 (2011).

20. Society, T. E. Instrumental Variables Regression with Weak Instruments Author ( s ): Douglas Staiger and James H . Stock Reviewed work ( s ): Published by : The Econometric Society Stable URL : http://www.jstor.org/stable/2171753 . *Society* **65**, 557–586 (2011).

21. Sanderson, E., Spiller, W. & Bowden, J. Testing and Correcting for Weak and Pleiotropic Instruments in Two-Sample Multivariable Mendelian Randomisation. *bioRxiv* 2020.04.02.021980 (2020) doi:10.1101/2020.04.02.021980.

22. Bowden, J., Davey Smith, G. & Burgess, S. Mendelian randomization with invalid instruments: effect estimation and bias detection through Egger regression. *Int. J. Epidemiol.* **44**, 512–525 (2015).

23. Bowden, J., Davey Smith, G., Haycock, P. C. & Burgess, S. Consistent Estimation in Mendelian Randomization with Some Invalid Instruments Using a Weighted Median Estimator. *Genet. Epidemiol.* **40**, 304–314 (2016).

24. Hartwig, F. P., Davey Smith, G. & Bowden, J. Robust inference in summary data Mendelian randomization via the zero modal pleiotropy assumption. *Int. J. Epidemiol.* **46**, 1985–1998 (2017).

25. Hemani, G., Tilling, K. & Davey Smith, G. Orienting the causal relationship between imprecisely measured traits using GWAS summary data. *PLOS Genet.* **13**, e1007081 (2017).

UOB Open
